# The first three months of the COVID-19 epidemic: Epidemiological evidence for two separate strains of SARS-CoV-2 viruses spreading and implications for prevention strategies

**DOI:** 10.1101/2020.03.28.20036715

**Authors:** Knut M. Wittkowski

## Abstract

About one month after the COVID-19 epidemic peaked in Mainland China and SARS-CoV-2 migrated to Europe and then the U.S., the epidemiological data begin to provide important insights into the risks associated with the disease and the effectiveness of intervention strategies such as travel restrictions and lockdowns (“social distancing”). Respiratory diseases, including the 2003 SARS epidemic, remain only about two months in any given population, although peak incidence and lethality can vary. The epidemiological data suggest that at least two strains of the 2020 SARS-CoV-2 virus have evolved during its migration from Mainland China to Europe. South Korea, Iran, Italy, and Italy’s neighbors were hit by the more dangerous “SKII” variant. While the epidemic in continental Asia is about to end, and in Europe about to level off, the more recent epidemic in the younger US population is still increasing, albeit not exponentially anymore. The peak level will likely depend on which of the strains has entered the U.S. first. The same models that help us to understand the epidemic also help us to choose prevention strategies. Containment of high-risk people, like the elderly, and reducing disease severity, either by vaccination or by early treatment of complications, is the best strategy against a respiratory virus disease. Lockdowns can be effective during the month following the peak incidence in infections, when the exponential increase of cases ends. Earlier containment of low-risk people merely prolongs the time the virus needs to circulate until the incidence is high enough to initiate “herd immunity”. Later containment is not helpful, unless to prevent a rebound if containment started too early.

**About the Author:** Dr. Wittkowski received his PhD in computer science from the University of Stuttgart and his ScD (Habilitation) in Medical Biometry from the Eberhard-Karls-University Tübingen, both Germany. He worked for 15 years with Klaus Dietz, a leading epidemiologist who coined the term “reproduction number”, on the Epidemiology of HIV before heading for 20 years the Department of Biostatistics, Epidemiology, and Research Design at The Rockefeller University, New York. Dr. Wittkowski is currently the CEO of ASDERA LLC, a company discovering novel interventions against complex (incl. coronavirus) diseases from data of genome-wide association studies.

## Introduction

The first cases of a new coronavirus strain, termed SARS-CoV-2 (<u>S</u>evere <u>A</u>cute <u>R</u>espiratory <u>S</u>yndrome <u>Co</u>rona<u>V</u>irus) by the International Committee on Taxonomy of Viruses (ICTV) ^(Cascella 2020)^, were reported on 31-12-2019 in Wuhan, the capital of the Hubei province of China.^(Jernigan 2020)^ As of 2020-04-25 10:00 CET, 2,744,744 symptomatic cases and 195,387 deaths have been reported from virtually every country in the northern hemisphere (see Section Data), The disease was termed COVID-19 by the WHO on 2020-02-11, and categorized as a pandemic on 2020-03-12, yet the details of the spread and their implications for prevention have not been discussed in sufficient detail.

As the virus moved from China via Europe to the U.S., several administrations have imposed “lockdown” restrictions.

For most of the first three months of the epidemic, much of the response was driven by “fear, stigma, or discrimination”^(Ren 2020)^, including naming SARS-CoV-2 the “China virus”^(Rogers 2020)^, despite the fact that seasonal respiratory zoonotic pathogens typically originate in China, where life-animal markets provide chances for animal viruses to transmit to humans.^(Malik 2020)^ On 03-23, the Veterans Health Administration issued a COVID-19 Response Plan envisioning a pandemic that would “last 18 months or longer and could include multiple waves of illness”.^(Stone 2020)^

Between 02-14 and 03-16, the Dow Jones fell 31% from 29,440 to 20,188, raising fears for the economy, in general, and retirement savings, in particular. By the end on 03-20, the Dow Jones was down at 19,173 (35%) from 02-14. On 03-26, the U.S. Senate approved a $2T stimulus package.

After three months, the epidemic has peaked in Asia, Europe, Oceania, and the Americas (Fig 1) and sufficient data are available to discuss important epidemiological characteristics of COVID-19 and the potential impact of interventions.

**Fig 1:**
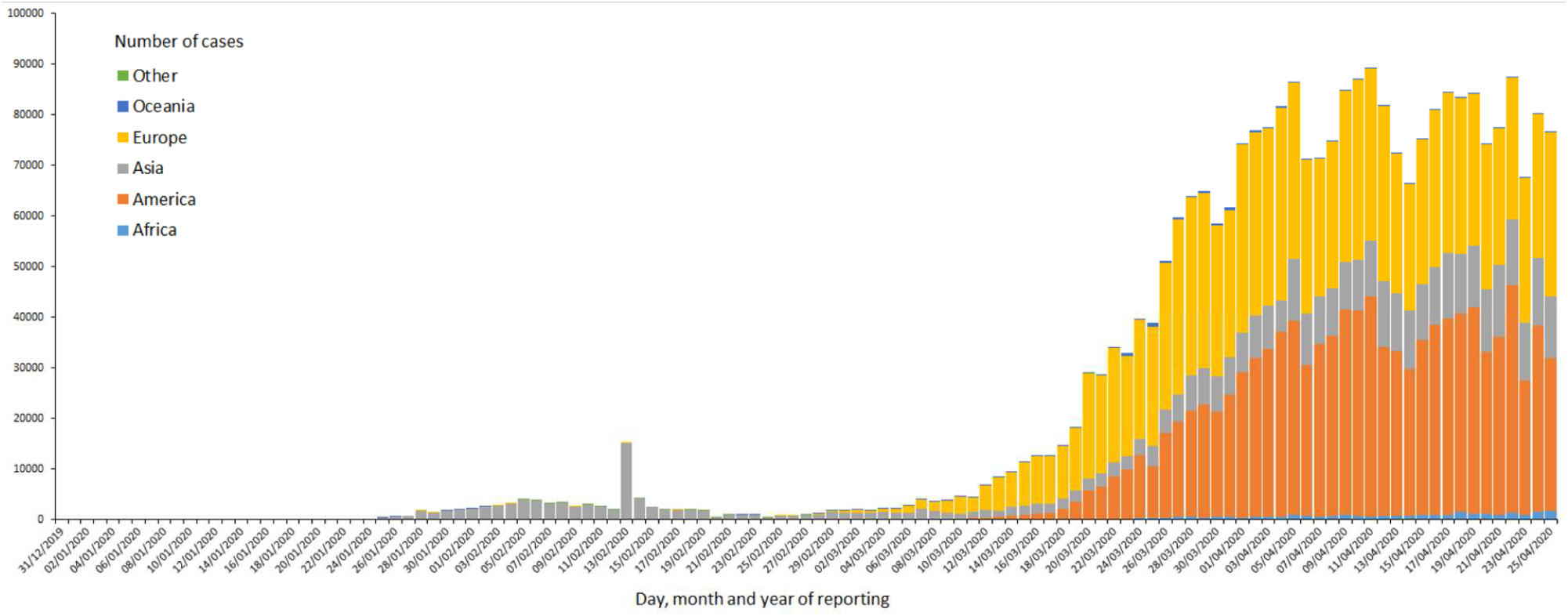
New Case Reports by Continent. ^(https://www.ecdc.europa.eu/sites/default/files/styles/is_full/public/images/novel-coronavirus-cases-worldwide-7-april-2020.png^) Note that the reports often do not distinguish between disease from SARS-CoV-2 and disease with detectable levels of SARS-CoV-2.

Changes in number of deaths follow the changes in number cases (albeit at a lower level) by about 1–2 weeks (Fig 2). During this season’s pandemic cases seem to have peaked in early April and deaths in mid-April.

**Fig 2:**
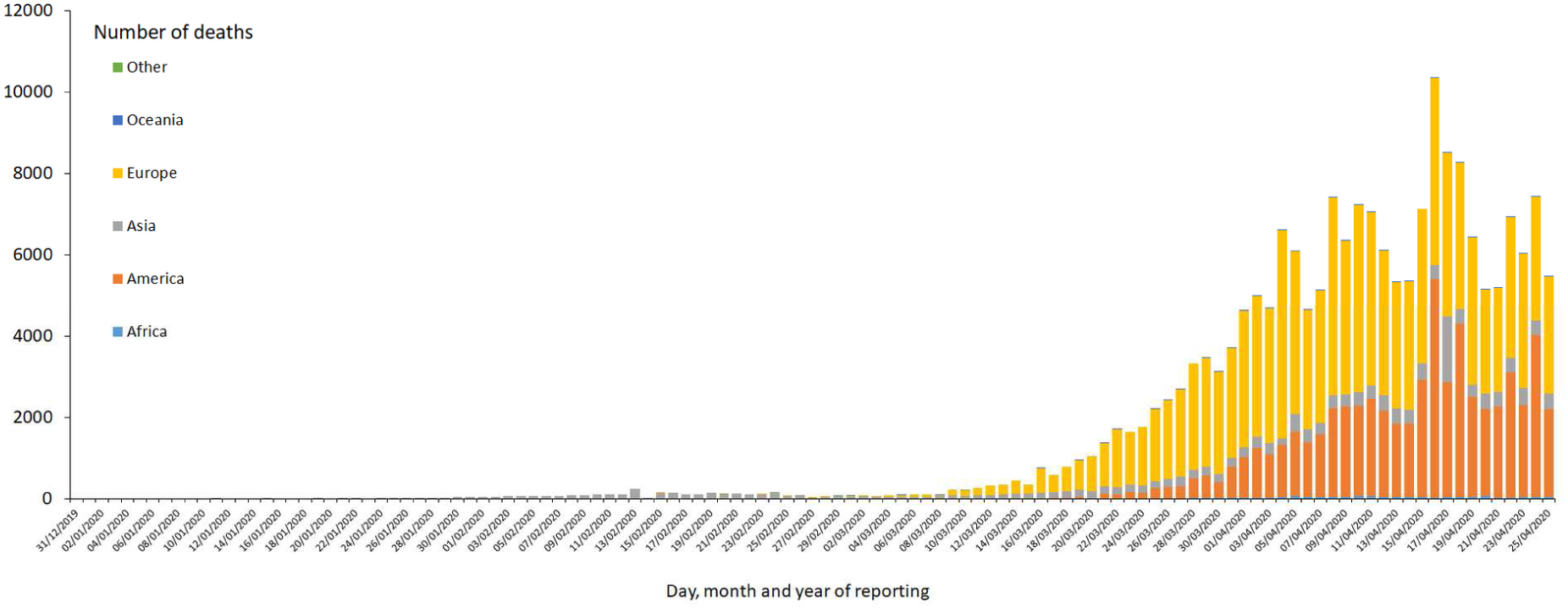
Death Reports by Continent. ^(https://www.ecdc.europa.eu/sites/default/files/styles/is_full/public/images/novel-coronavirus-cases-worldwide-deaths-7-april-2020.png)^ Note that the reports often do not distinguish between death from SARS-CoV-2 and death with detectable levels of SARS-CoV-2.

As this year’s seasonal flu season ends in the northern hemisphere (unless there should be an outbreak in maritime Asia, India, or Russia), we can discuss both the infectiousness and the lethality of the virus, two important characteristics to assess public health impact of the disease and the implication for prevention strategies. An important finding is that the interventions in several countries started too early (prolonging the time the virus stayed in the population and, potentially, increasing the number of deaths) or too late (being ineffective). Hence, the timepoint when a public health intervention starts during the course of the epidemic (especially the “turning point” where the increase in new cases begins to decline) is crucial for the impact of the intervention.

## Materials and Methods

### Data

All data (with the exception of NYC data) were downloaded on 2020-04-16 from the European Centre for Disease Prevention and Control (ECDC) Web site at https://www.ecdc.europa.eu/en/publications-data/download-todays-data-geographic-distribution-covid-19-cases-worldwide, where data are collected daily between 6:00 and 10:00 CET. Updates were collected from the Johns Hopkins online tracker available at https://systems.jhu.edu/research/public-health/ncov/. New York City data was downloaded from https://www1.nyc.gov/assets/doh/down-loads/pdf/imm/covid-19-daily-data-summary.pdf.

Population data were accessed from https://www.worldometers.info/world-population/population-by-country/ on 2020-03-12. Data on ages by country were accessed from https://data.worldbank.org/indicator/SP.POP.65UP.TO.ZS.

## Methods and models

### Statistical and Bioinformatics Methods

The data were processed with MS Excel. To avoid biases from inappropriate model assumptions only basic descriptive statistics were employed. In some cases, data from only the day before or after (or both) was averaged (up to a three day moving average) to reduce the effects of apparent reporting artifacts (Darwin’s *natura non facit saltum*,^(Berry 1985)^) without creating undue biases. The two “smoothers” applied were:

- averaging x_0_ with a previous x_−1_ (or, rarely, following x_+1_) data and
- applying a moving average of (x_− 1_, x_0_, x_+1_)→(2 x→_1_ + x_0_, x→_1_ + x_0_ + x_+1_, x_0_ +2 x_+1_)/3 No other changes were applied to the data.

Like China in mid-February, the German Robert-Koch-Institute (RKI) changed the reporting system in mid-March, which resulted in a near 6-fold increase of the data reported on 03-20 over the data reported on 03-19. To account for such isolated event, the data of this and several previous days was averaged (as indicated in the Figures) under the assumption that those changes reflect cases that had previously been diagnosed, but were not reported.

To guide with visual impression, moving averages of the 4 days leading up to the current day were added to the Figures.

### Epidemiological Models

If a disease causes immunity after an infectious period of a few days only, like respiratory diseases, an epidemic extinguishes itself as the proportion of immune people increases. Under the SIR model,^(Kermack 1991)^ for a reproduction number^(Dietz 1993)^ (secondary infections by direct contact in a susceptible population) of R_0_=1.5–2.5 over 7 days (recovery rate: γ=1/7=.14), the noticeable part of the epidemic lasts about 90–45 days (R_0_/γ=β=.21–.36) in a homogeneous population of 10M. The period is shorter for smaller more homogenous and longer for larger, more heterogenous populations. For a given infectious period 1/γ (here, e.g., 7 days. SARS and COVID-19 incubation period plus 2 days^(Lauer 2020)^), R_0_ also determines how long it will take for early cases to become visible after a single import (150–60 days), the peak prevalence of infections (5–22%), and how many people will become immune (55–90%). To allow for comparisons between models, an arbitrary proportion of symptomatic cases among those becoming infected (.05%) is used and 2% of cases are assumed to die.

The model reflects that the key milestones of the epidemic, the turning point/half maximal point in infections (red, day 68), the turning point/half maximal point in the number of cases (day 75, orange), the peak in infections (day 83, red), the peak in cases (day 70, orange) and the peak in deaths (day 77, gray) follow each other, following the previous milestone after about a week.

## Results

### Time-course by country/region

#### Incidence by Country. Norther Hemisphere

Table 1 shows the raw daily incidence by population sizes for countries with epidemiological relevance in the northern hemisphere. Countries within proximity are grouped by their peak incidence (red background).

**Table 1:**
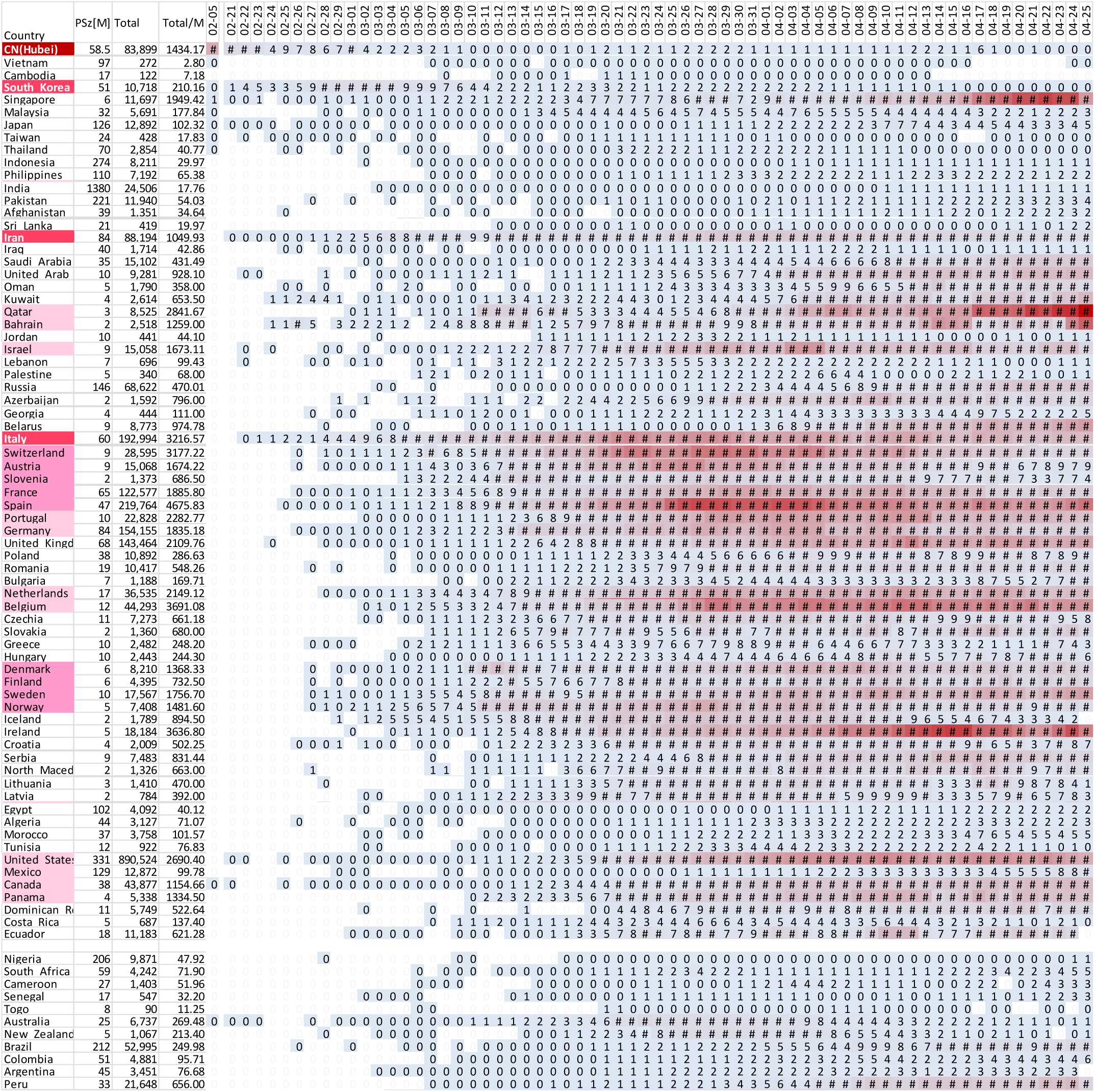
Incidence by Country. Dates: Feb 13 (peak intensity in Mainland China, mostly Hubei and neighboring provinces), Feb 19 to Mar 22. Countries with low population size (PSz) or low number of cases (Total) are hidden. Red background indicates countries/dates with high incidence per capita. Countries/regions are grouped by proximity among each other and distance from Mainland China.

**Table 2:** Epidemiological Timepoints by Country. Top: 2003 SARS, Bottom: 2020 SARS-CoV-2

|  | First cases | Gap [d] | Begin (>.1/1M) | Gap [d] | Peak | Gap [d] | End (<.1/1M) | Begin to End [d] |  |
| --- | --- | --- | --- | --- | --- | --- | --- | --- | --- |
| CA/SGH | 02-23 |  | 03-02 | 18 | 03-20 | 28 | 04-18 | 28 | (Svoboda 2004) |
| CA/NYG | 04-20 |  | 04-27 | 30 | 05-27 | 6 | 06-03 | 36 | (Svoboda 2004) |
| Guangdong | 2002 | >50 | 01-19 | 22 | 02-11 | 81 | 05-02 | 103 | (Cao 2019) |
| Shanxi |  |  | 03-15 | 34 | 04-19 | 30 | 05-19 | 64 | (Cao 2019) |
| Beijing |  |  | 03-05 | 49 | 04-24 | 30 | 05-24 | 69 | (Zhou 2003; Cao 2019) |
| Mongolia |  |  | 03-22 | 22 | 04-14 | 40 | 05-24 | 62 | (Cao 2019) |
| Hebei |  |  | 04-04 | 20 | 04-24 | 24 | 05-19 | 44 | (Cao 2019) |
| Tianjin |  |  | 04-14 | 8 | 04-22 | 12 | 05-04 | 20 | (Cao 2019) |
| HK/PWH | 02-21 |  | 03-11 | 6 | 03-17 |  |  |  |  |
| HK/AG |  |  | 03-20 | 4 | 03-24 | 29 | 04-13 |  | (Zhou 2003; Leung 2004) |
| HK/ |  |  |  |  | 04-12 | 54 | 06-06 |  |  |
| Taiwan | 02-24 | 23 | 03-17 | 38 | 04-25 | 50 | 06-14 | 88 | (Small 2003; Yeh 2004) |
| Singapore |  |  | 03-15 | 20 | 04-05 | 24 | 04-29 | 44 | (Small 2003; Goh 2006) |
| Vietnam |  |  | 02-23 | 11 | 03-04 | 28 | 04-06 | 39 | (Shi 2003) |
| Median 2003 |  |  | 03-15 | 20 | 04-13 | 29 | 05-19 | 44 |  |
| China (Hubei) | 2019 | >50 | 01-17 | 18 | 02-05 | 43 | 03-18 | 61 | 0 in Hubei |
| S- Korea | 01-20 | 31 | 02-19 | 12 | 03-01 | 45 | 04-15 | 57 | Ongoing low level |
| Iran | 02-20 | 2 | 02-22 | 14 | 03-07 |  |  |  | Several waves? |
| Italy | 01-31 | 22 | 02-22 | 30 | 03-22 |  |  |  | 2<sup>nd</sup> wave? |
| Germany | 01-28 | 30 | 02-27 | 36 | 04-03 |  |  |  | Reporting system |
| France | 01-25 | 31 | 02-27 | 34 | 03-31 |  |  |  |  |
| US | 01-25 | 40 | 03-05 | 34 | 04-09 |  |  |  |  |
| Median 2020 |  | 31 | 02-22 | 30 | 03-22 |  |  |  |  |

It should be noted, however, that there is no uniform definition of “cases”. In some countries a case needs to have symptoms, in other countries, it suffices to have antibodies (be immune).

Among the Hubei population of 58.5M, the incidence rose from the first case reported in late 2019 to about 60 new cases per million people per day by 02-05 and then steadily declined (Fig 4) from ∼4000 on 02-05 to below 50 cases per day since 03-08. Since 03-31 no cases were reported in Vietnam and only isolated cases in Cambodia. The cumulative incidence was about 1,400/M.

**Fig 3:**
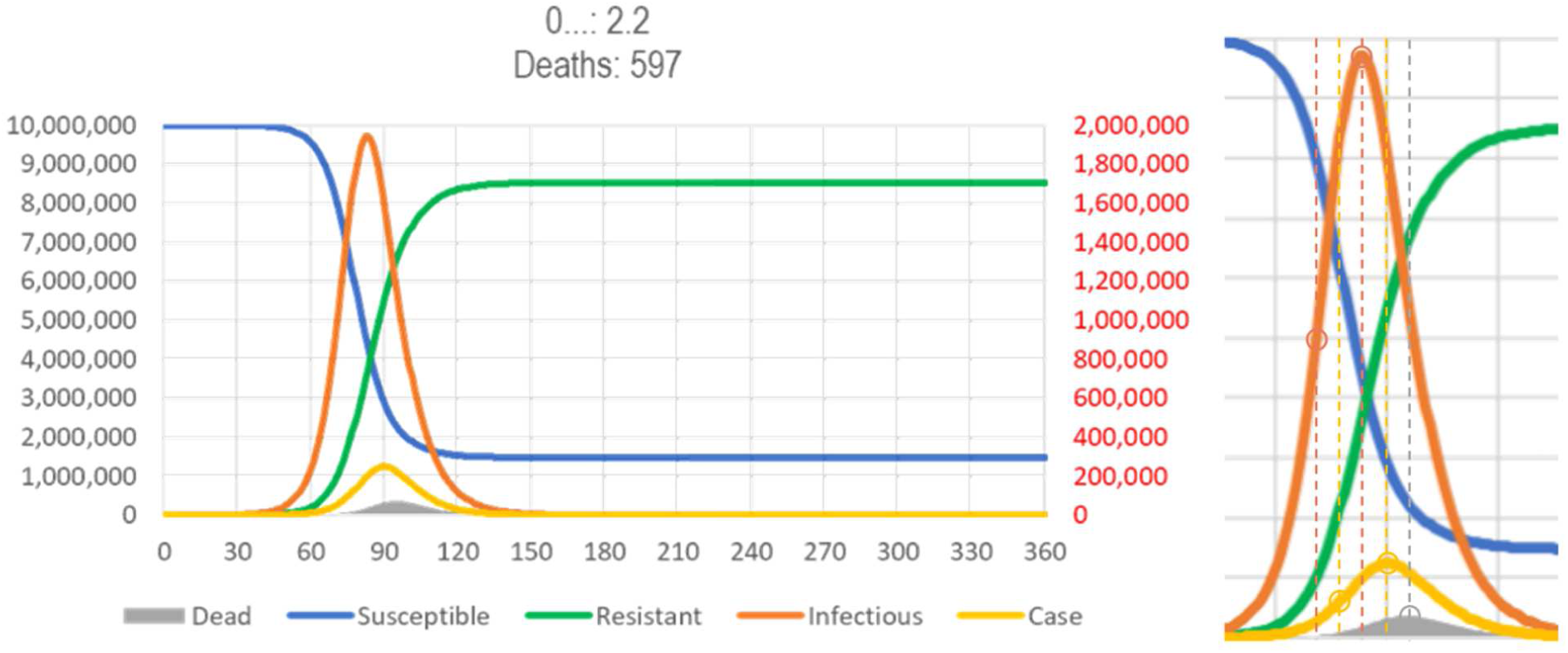
SIR Model of SARS. Number of susceptible (blue), infectious (red), resistant (green), case (orange) and dead (gray) people after a population of 10,000,000 susceptible people is exposed to 20 subjects infected carrying a novel virus. Assumptions: R_0_ = 2.2, infectious period = 7 days,^(available from https://app.box.com/s/pa446z1csxcvfksgi13oohjm3bjg86ql)^

**Fig 4:**
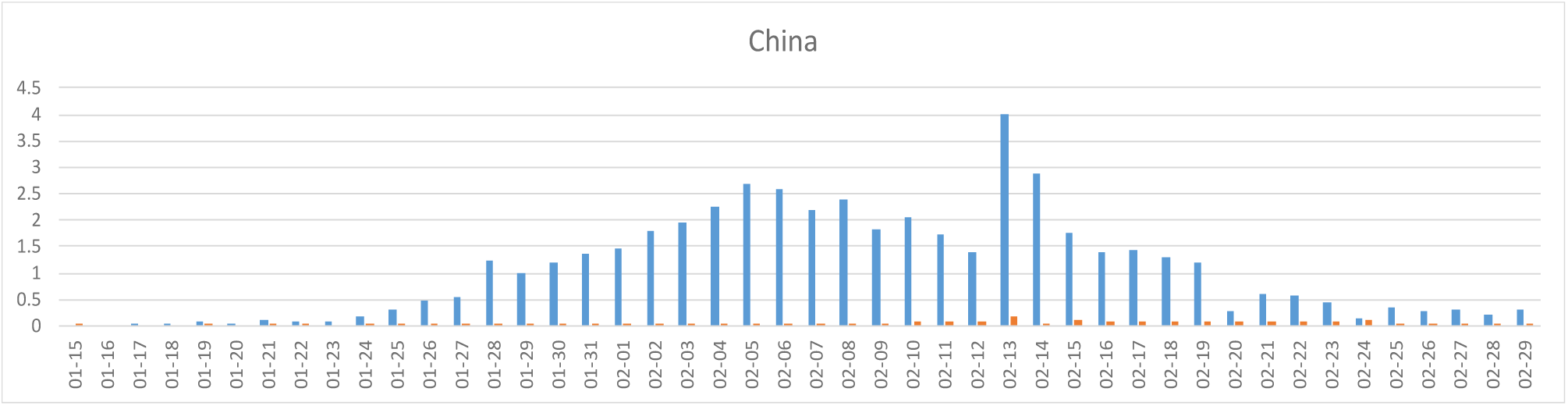
COVID-19 cases in Mainland China. Blue: cases/M/d, red: deaths/M/d. Around Feb 13, the case definition was expanded, resulting in additional cases from previous days being added. Hence, the 02-13 cases have been truncated. Most cases were seen in the Hubei province of 58.5M people (see Table 1 for population sizes).

In continental South Korea (population 51M, cumulative incidence 210/M), the incidence rose to a peak of about 14/M/d between 02-29 and 03-02, before declining to continuing low rate of less than 150 cases per day (∼2/M/d) since 03-12 (Fig 5a).

**Fig 5:**
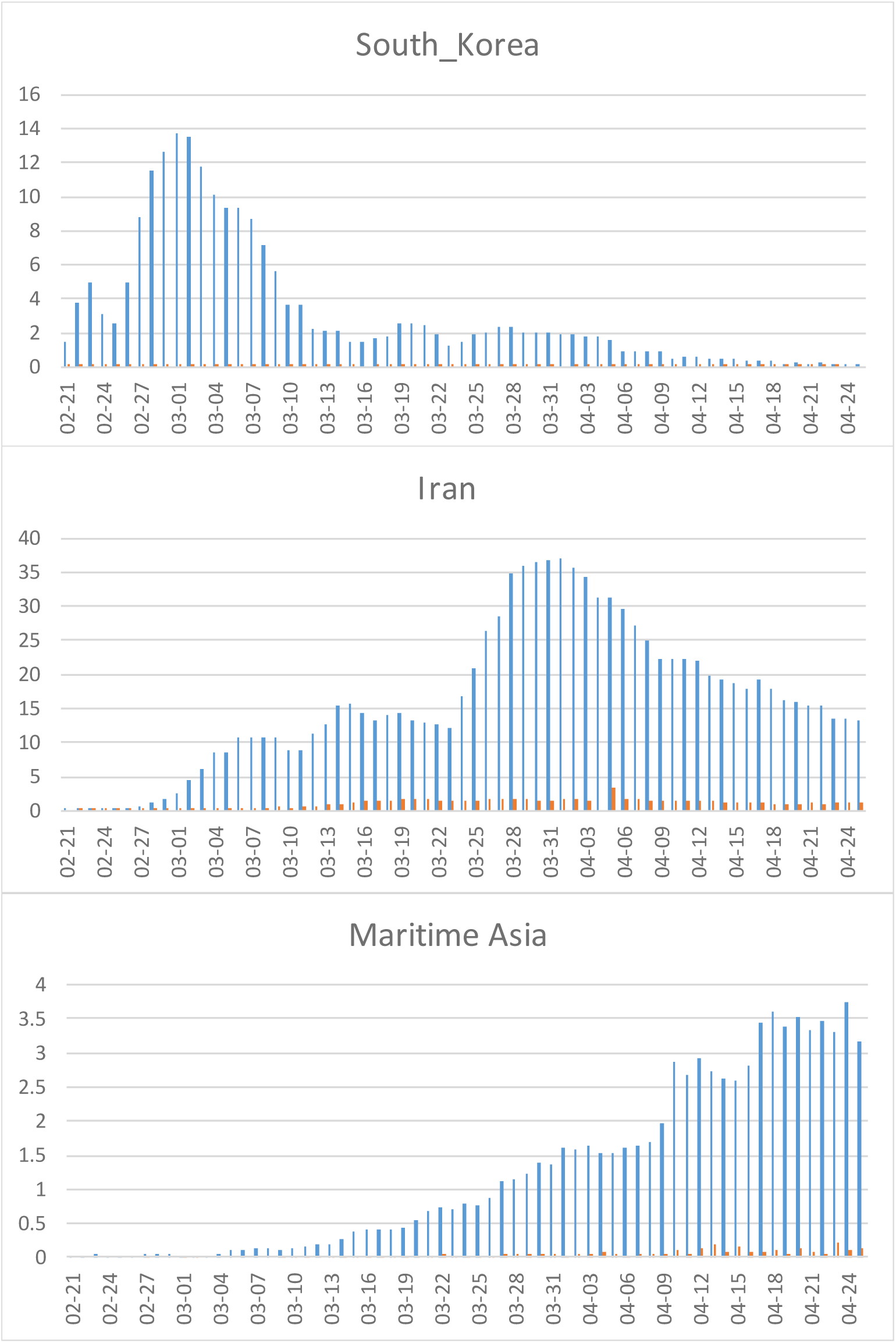
COVID-19 cases in Asia. See Fig 1 for legend.

In Iran (cumulative incidence 1050/M), incidence began to rise about a week after South Korea. The top incidence before a new wave started on 03-23 (∼15.5 cases/M/d) was about the same. The increase thereafter with a peak around 04-01 at 37M/d may indicate a “rebound” into a population not sufficiently immunized by the previous wave(s). Lethality in Iran was notably higher than in South Korea and followed the increase in cases (see the SIR model, above) with a delay of several days (Fig 5b).

By Mid-January 2020, the first cases of COVID-19 were seen in other Asian countries, but incidence remained below about 3/M/d outside of continental China (Fig 5c); only Malaysia/Brunei and Singapore did the incidence raise above 5/M/d; in Singapore incidence has recently increased to 185/M/d. In Japan with the largest proportion of people >65 years of age in the world (28%) remained below 1.1/M/d until recently, when incidence increased to ∼7/M/d. Hence, Japan may be increasing to the level seen in South Korea, unless spread of the virus ends with the end of the flu season, as indicated in a drop over the last few days.

From 03-19 to 03-20, several European countries have seen a more than two-fold increase in the number of cases reported (Germany: 570%, San Marino: 340%, Ireland: 260%, Switzerland: 240%, Austria: 202%). As *natura non facit saltum* (Darwin: nature doesn’t jump),^(Berry 1985)^ such abrupt increases must be, at least in part, the result of reporting or other artifacts. In Germany, for instance, the reporting system was changed between 03-16 and 03-19, so that the number likely includes cases previously reported only through a parallel system. France, Italy, and Spain also reported an unusual increase by 27–35 percent. Since 3-26, all these countries reported the incidence to decline with a wave pattern consistent with weekday effects of reporting.

Cumulative incidence per capita in Italy’s neighbors Spain and Switzerland has exceeded that of Italy (Table 1, Fig 6a), at levels higher than in the Hubei province (all with a population of 50–60 M). Among other European countries, only the BeNeLux region reached similar levels (Table 1).

**Fig 6:**
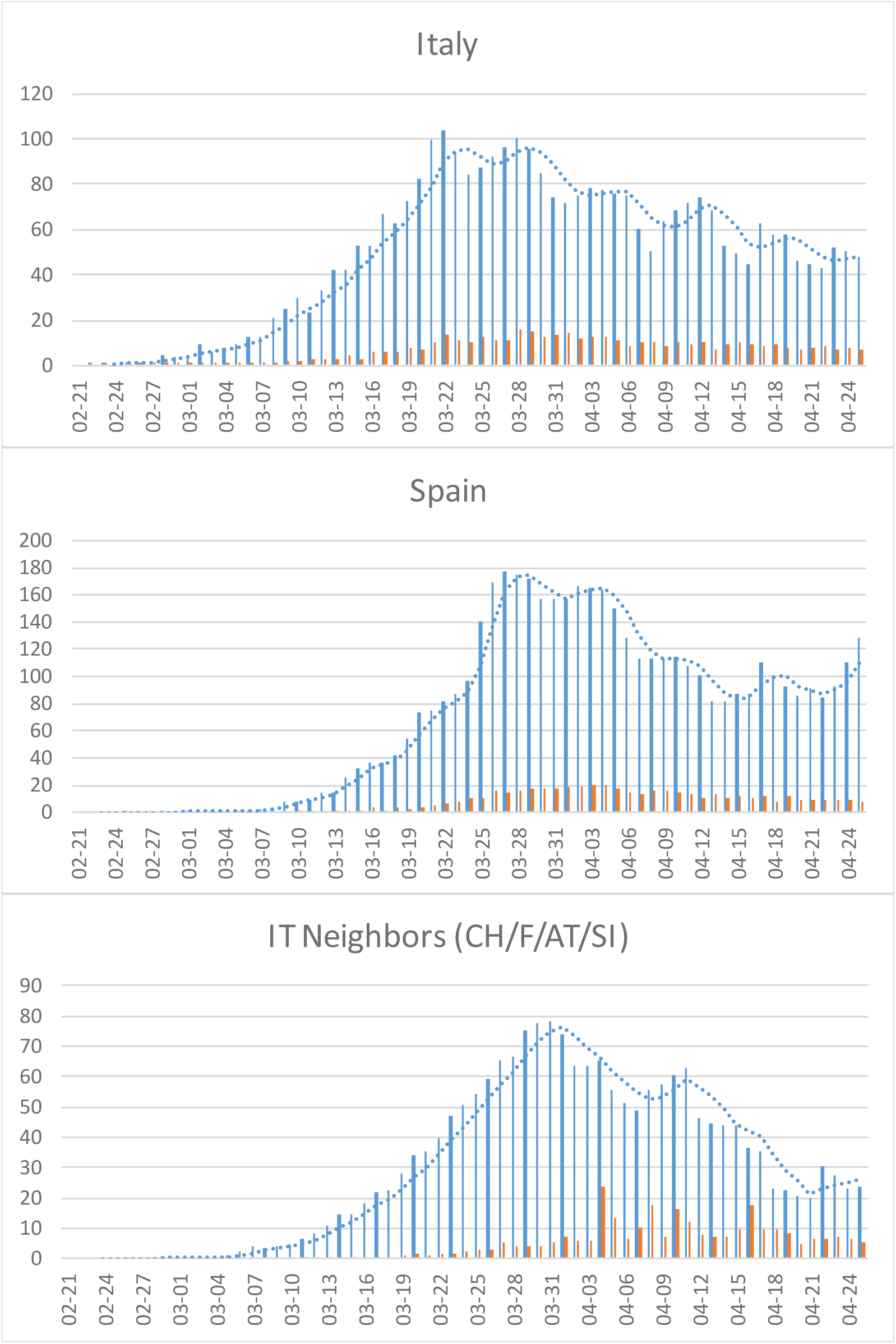
COVID-19 cases in Italy and it’s neighboring countries. Italy (top), Spain (center), European countries neighboring Italy. See Fig 1 for legend.

In Italy (Fig 6), incidence has been declining continuously from the 100/M/d peak around 03-22, after rising for about 4 weeks from 1/M (02-26..∼03-22). In Hubei and South Korea, instead incidence took only 2 weeks to peak (01-19..02-05, Fig 4, and 02-21..03-06, Fig 5) and then declined by at least 85% within another 2 weeks. The much slower decline in Italy (compared, e.g., to China and Hubei) is consistent with insufficient herd immunity having been developed before the peak incidence was reached (Fig 3). While Italy and its neighbors (Europe/high) peaked in 03-27/28 (Fig 7). Overall, the incidence in Europe reached its peak on 03-28 in the countries with high lethality and on 04-04 in the other countries.

**Fig 7:**
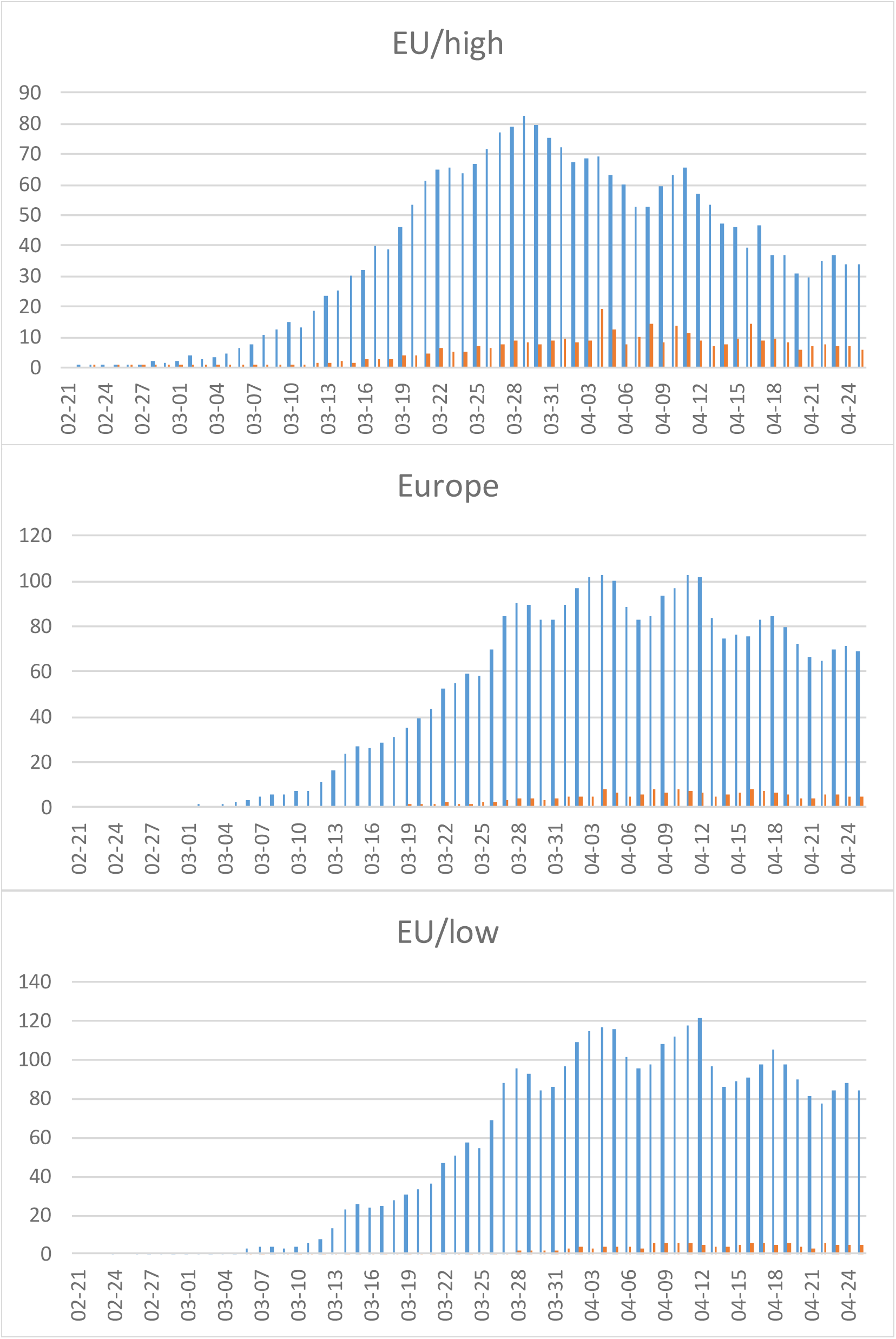
COVID-19 cases in Europe. Early onset/high lethality (IT and neighbors, top), total (middle), and late onset/low lethality (other European countries, bottom). See Fig 1 for legend.

Austria is notable for both its early peak and the steep decline; since infections predate cases by at least a week, SARS-CoV-2 in Austria may already have been eradicated.

Among the main Scandinavian countries, there was no difference in peak incidence (65/M/d, except for a recent spike in Sweden to 80) and a slower increase in incidence and a higher lethality in Sweden than in both Norway and Denmark (Fig 9).

**Fig 8:**
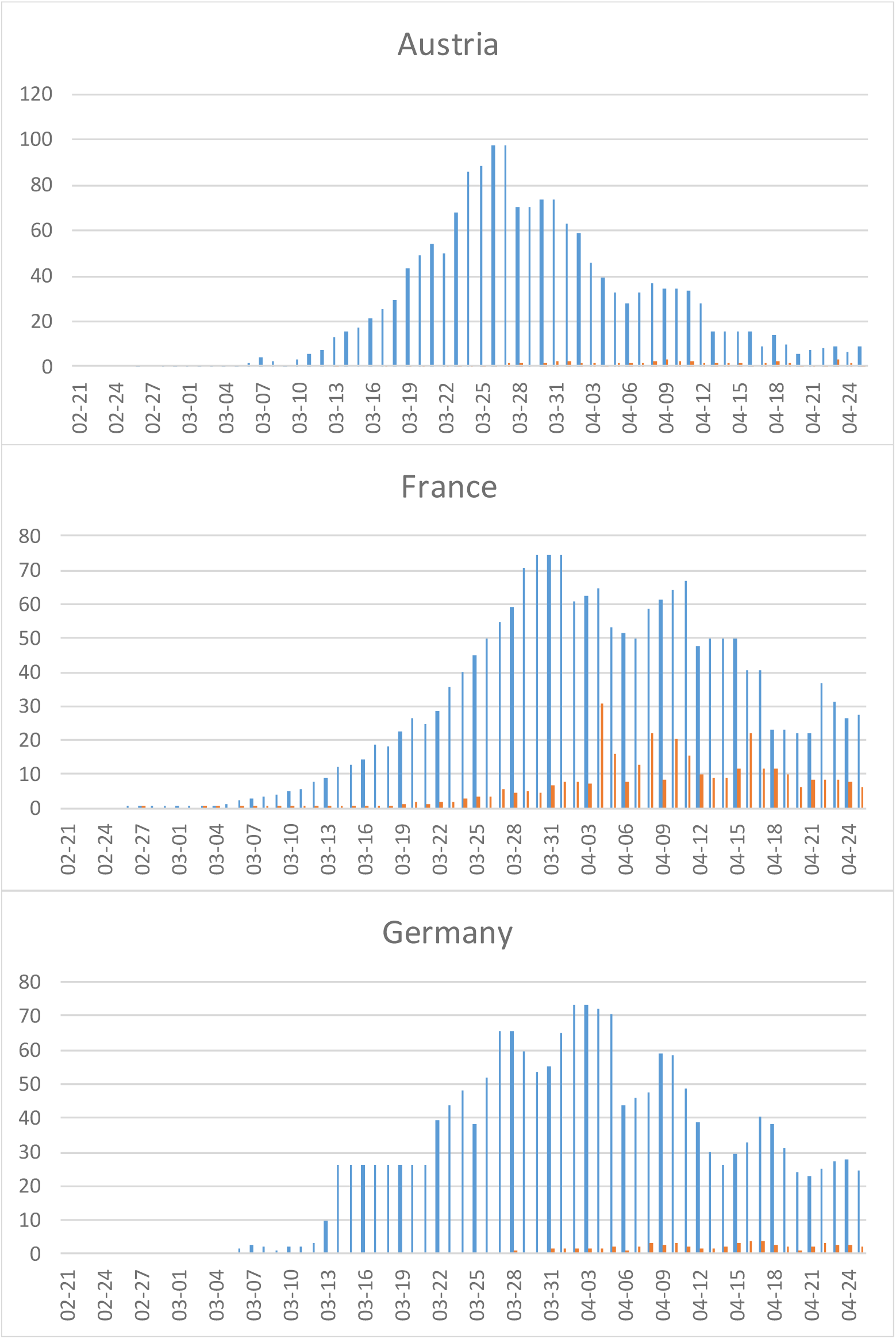
COVID-19 cases in selected European countries. See Fig 1 for legend. Data in Germany is based on cases reported electronically to the Robert-Koch-Institute (RKI) and transmitted to the ECDC, but the RKI also provides two sources of data on its Web site that are difficult to reconcile with these data. To account for a temporary peak in Germany on 03-20/21, when the reporting system was changed, the 03-14..03-21 were averaged. 6082 cases have been added for Germany based on ^(https://www.rki.de/DE/Content/InfAZ/N/Neuartiges_Coronavirus/Situationsberichte/2020-04-04-de.pdf?blob=publicationFile)^

**Fig 9:**
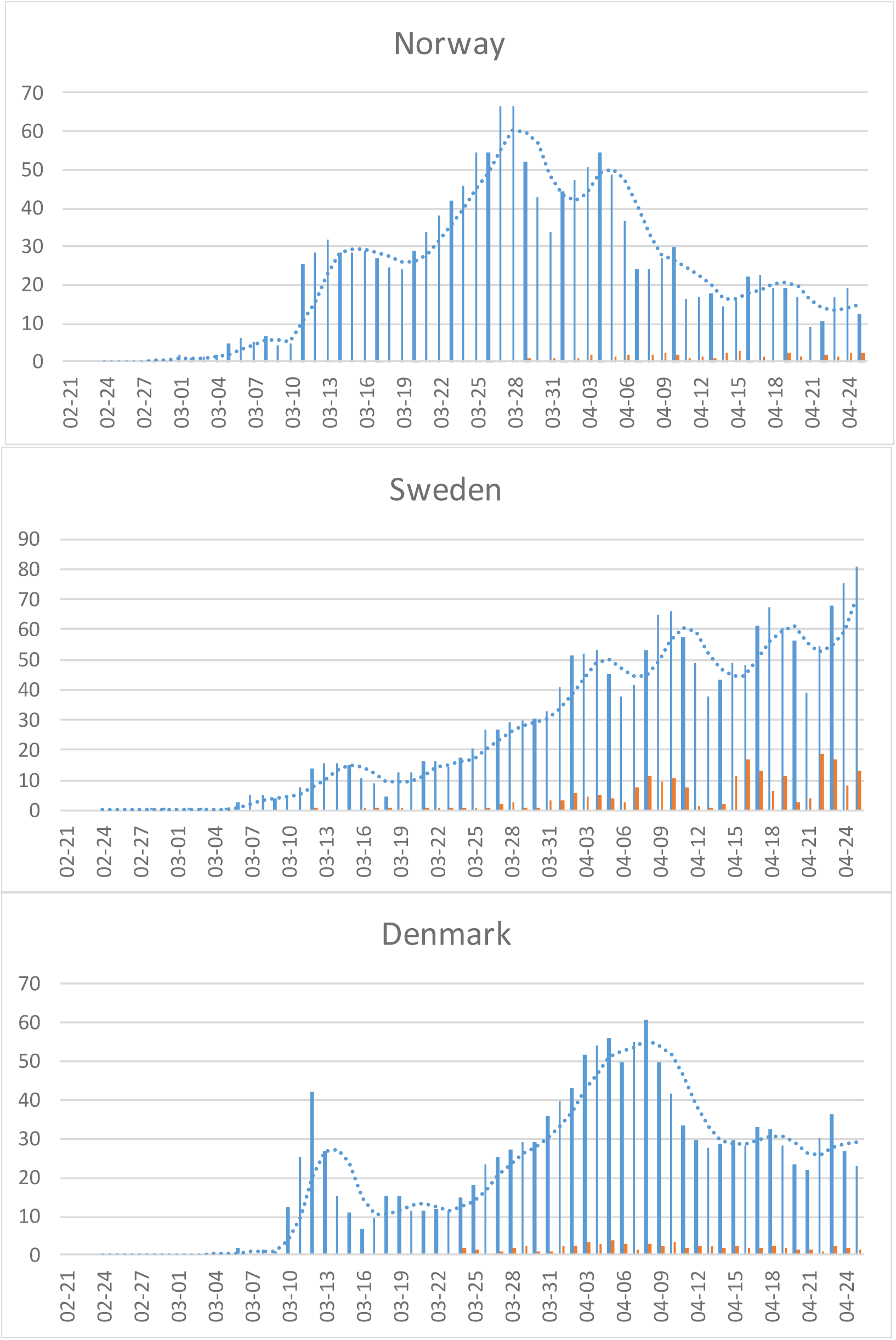
COVID-19 cases in the Scandinavia. See Fig 1 for legend.

The epidemic in North America started later, especially in the US and most cases were imported from Europe (except for a few isolated cases likely imported directly from Asia). The increase in incidence is consistent with the dynamics of an earlier epidemic and the cumulative incidence in the US (1841/M), still lower than Europe, as a whole (2426/M), and especially Italy (2708) or Spain (3671/M). The US reached the “turning point” (half the height of the 04-09/10 peak), where the rate of new cases begins to decline (when 50% of the peak incidence is reached) at about 03-27, a week later than Europe’s 03-21 (Fig 7). Levels in Canada remained below those in the U.S and are also declining. In New York City, which is particularly hard hit, the number of new cases seem to have peaked in early April and is also already declining. Lethality should follow suit with a delay of about a week.

A particular problem arises from changes of case definitions in the midst of an epidemic, as early on in China and now in the US.

### Incidence by Country. Southern Hemisphere

Most parts of the southern hemisphere (with the possible exception of Chile) have seen only few cases, but Oceania (Australia and New Zealand) shows evidence for an epidemic with a peak incidence on 03-26/27. As in Canada and, earlier, in Germany and other European countries, isolated spikes tend to reflect delayed reporting, rather than changes in incident trends. The number of deaths is extremely low (Fig 11).

**Fig 10:**
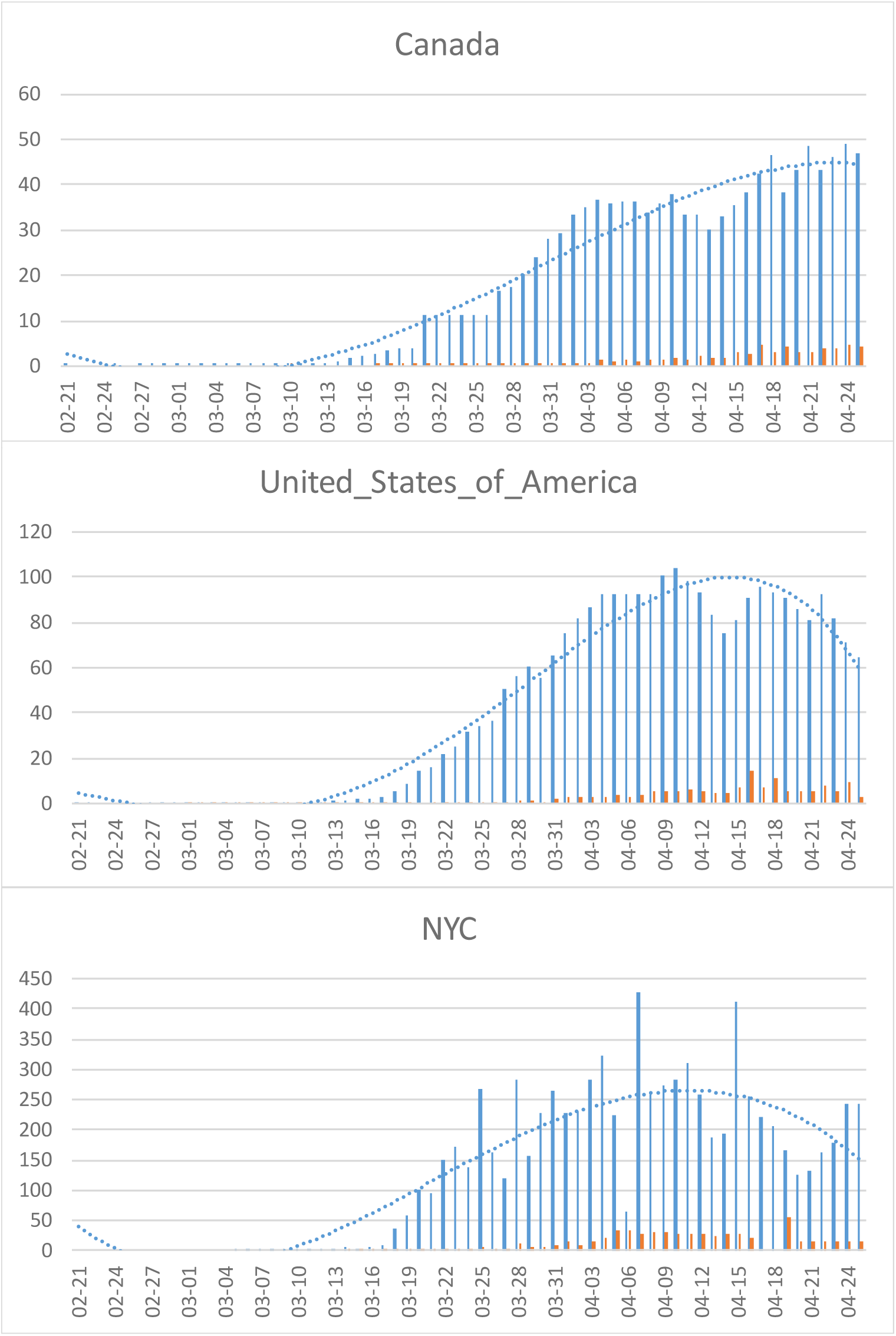
COVID-19 cases in the North America. See Fig 1 for legend. To account for an isolated peak in the Canadian data on 03-26, the data for 03-21..03-26 were averaged. The data in NYC are too irregular and incomplete to fit a moving average.

**Fig 11:**
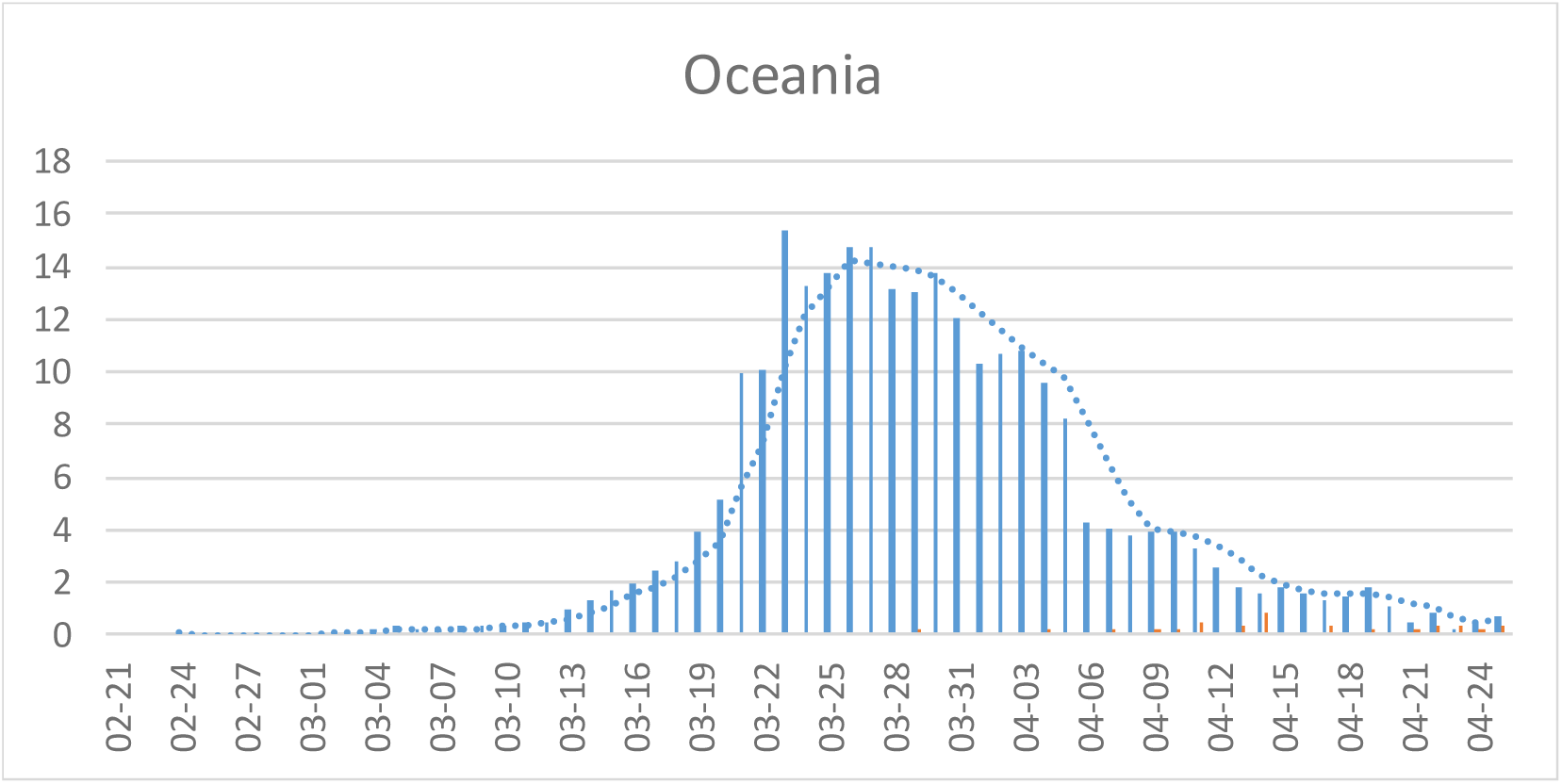
COVID-19 cases in the Oceania (Australia and New Zealand). See Fig 1 for legend.

### Modeling Results for Effectiveness of Containment (Lockdowns)

The effect of reducing the reproduction number by reducing the number of contacts (lockdowns) depends on when it starts in the course of the epidemic. Fig 12 shows the effect of a one-month intervention cutting R_0_ in half starting at the point of the peak prevalence of infectious subjects. Compared to Fig 3, the duration of the epidemic is shortened, albeit at the price of reducing the R/S ratio, so that a subsequent epidemic with the same or a similar virus (cross-immunity) could start earlier.

**Fig 12:**
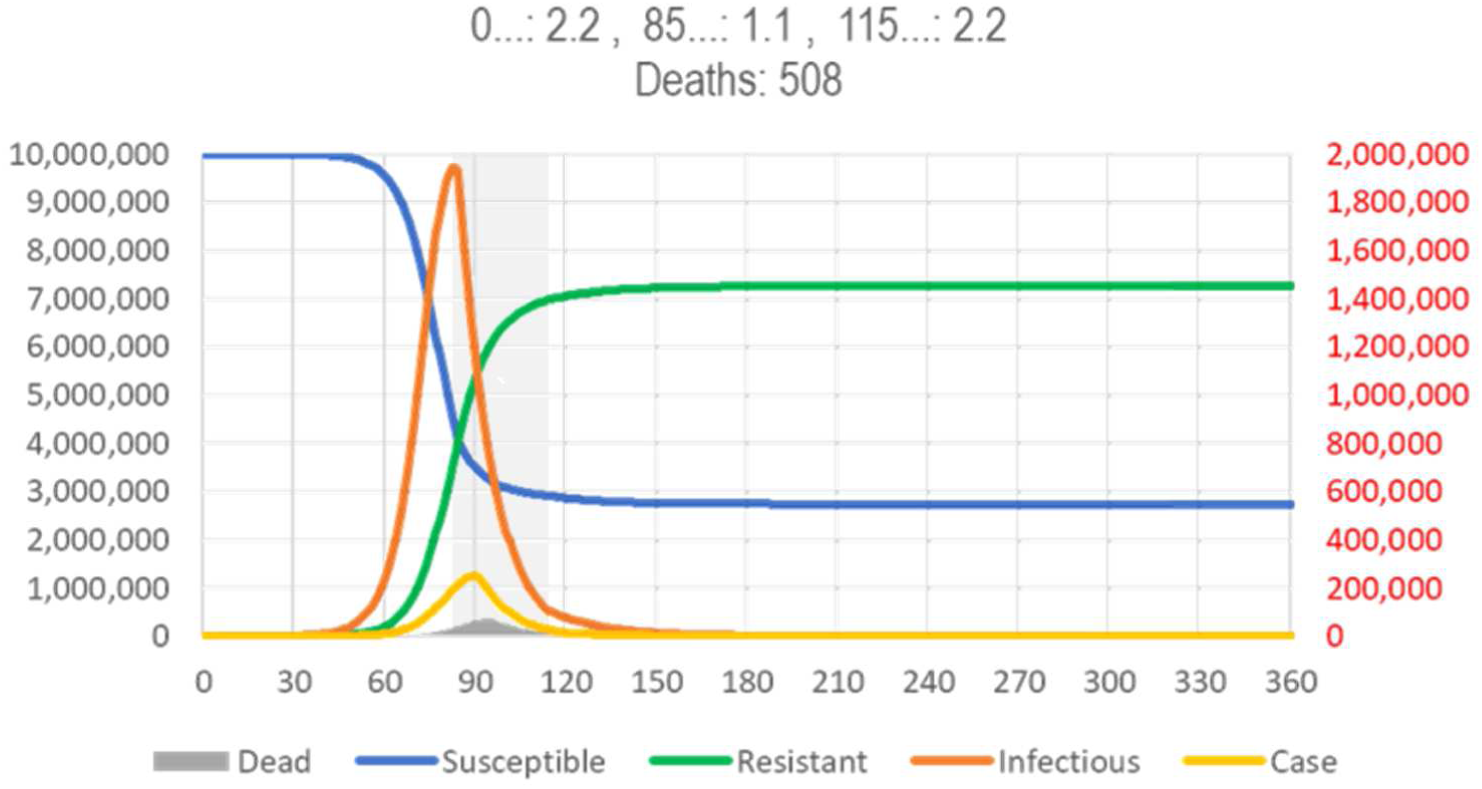
SIR Model of SARS, Window of Opportunity for Fast Eradication of the Epidemic. (see Fig 3 for legend). The gray area indicates the period where containment can give a “coup de grace” to a respiratory disease epidemic. The more narrow bell curve with a post-peak intervention indicates the reduction in number of infections and, thus, deaths^.(spreadsheet for model calculations available from https://app.box.com/s/pa446z1csxcvfksgi13oohjm3bjg86ql)^

Fig 13 shows a one-month intervention starting about two weeks earlier, at the turning point where the curve of the new cases changes from increasing faster to increasing more slowly. This intervention reduces the number of deaths, but the epidemic is extinguished two months later and the R/S ratio (“herd immunity”) is further decreased.

**Fig 13:**
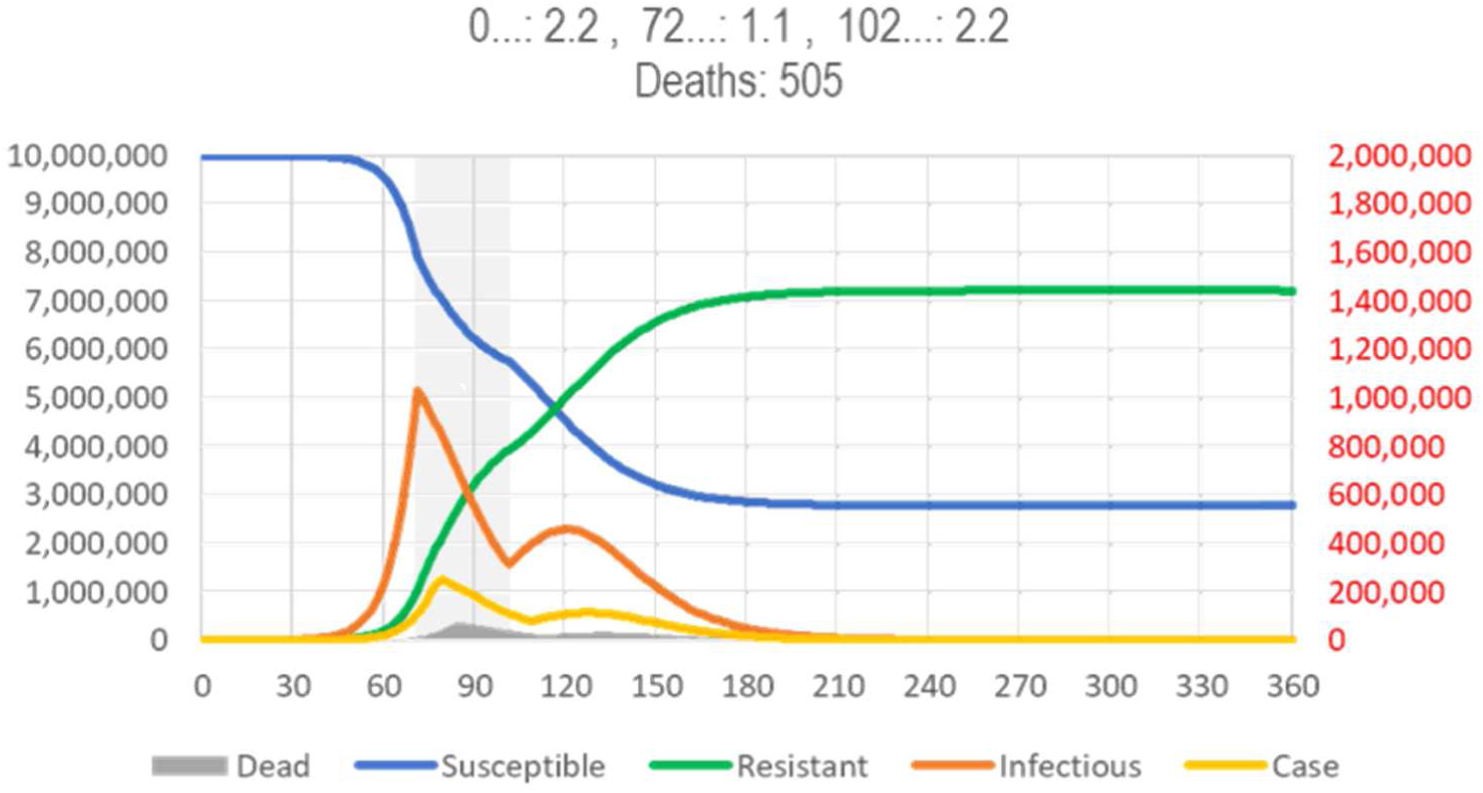
SIR Model of SARS, Window of Opportunity for Maximal Reduction of Total Deaths. (see Fig 3 for legend). The gray area indicates the period where containment can have the most impact on total number of deaths. However, the epidemic is not eradicated.^(spreadsheet for model calculations available from https://app.box.com/s/pa446z1csxcvfksgi13oohjm3bjg86ql)^

The SIR model is flexible enough to reflect a “phased in” intervention, which starts with a period of lower intensity (higher R_0_) to allow people to adjust, before rolling in a more restrictive intervention (higher R_0_). The Fig 14a shows the intervention of Fig 13 starting with a week of reduced intensity. Compared to Fig 13, reducing intensity during the period where herd immunity reduces the rebound, but – somewhat counterintuitively – also the number of deaths. Extending the duration of the intervention (almost) extinguish the disease (and further reduce number of deaths by 10%).

**Fig 14:**
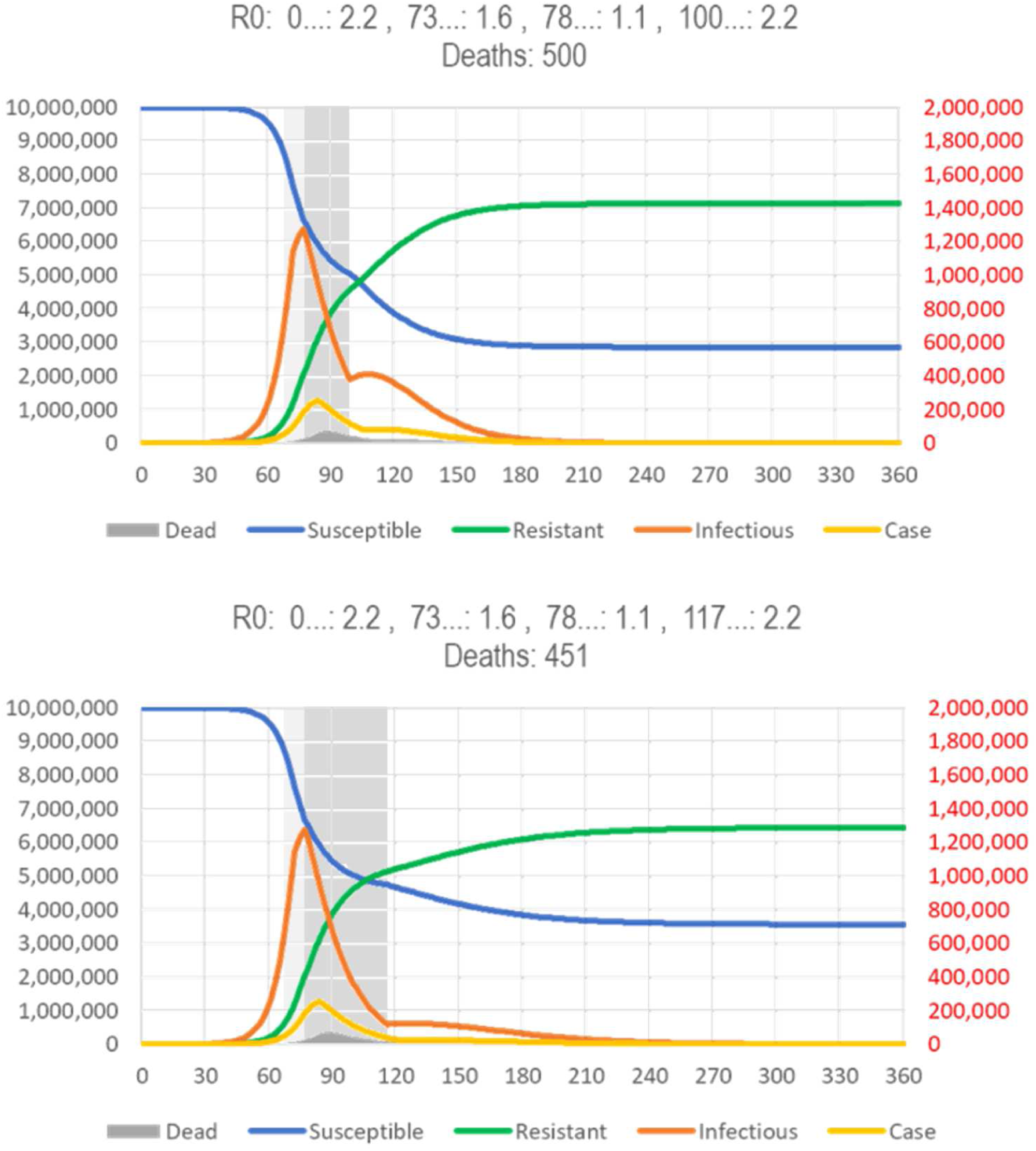
SIR Model of SARS, Phased in Restrictions. (see Fig 3 for legend). The gray areas indicate the periods of low (5 d) and high intensity (22/40 d) restrictions)

Fig 15 shows the detrimental effect of an intervention that starts even earlier, about two weeks before the turning point (Fig 13). Even if the intervention is extended from one to four months, no herd immunity is created and, thus, the epidemic rebounds and will run eight months, instead of three (Fig 3) or less (Fig 12). To avoid or even reduce the rebound, one would have to end the restrictions in the lowest risk populations (school children, young adults) first to increase the immune/susceptible ratio (the effects of targeting subpopulations differently are not accounted for in simple SIR models).

**Fig 15:**
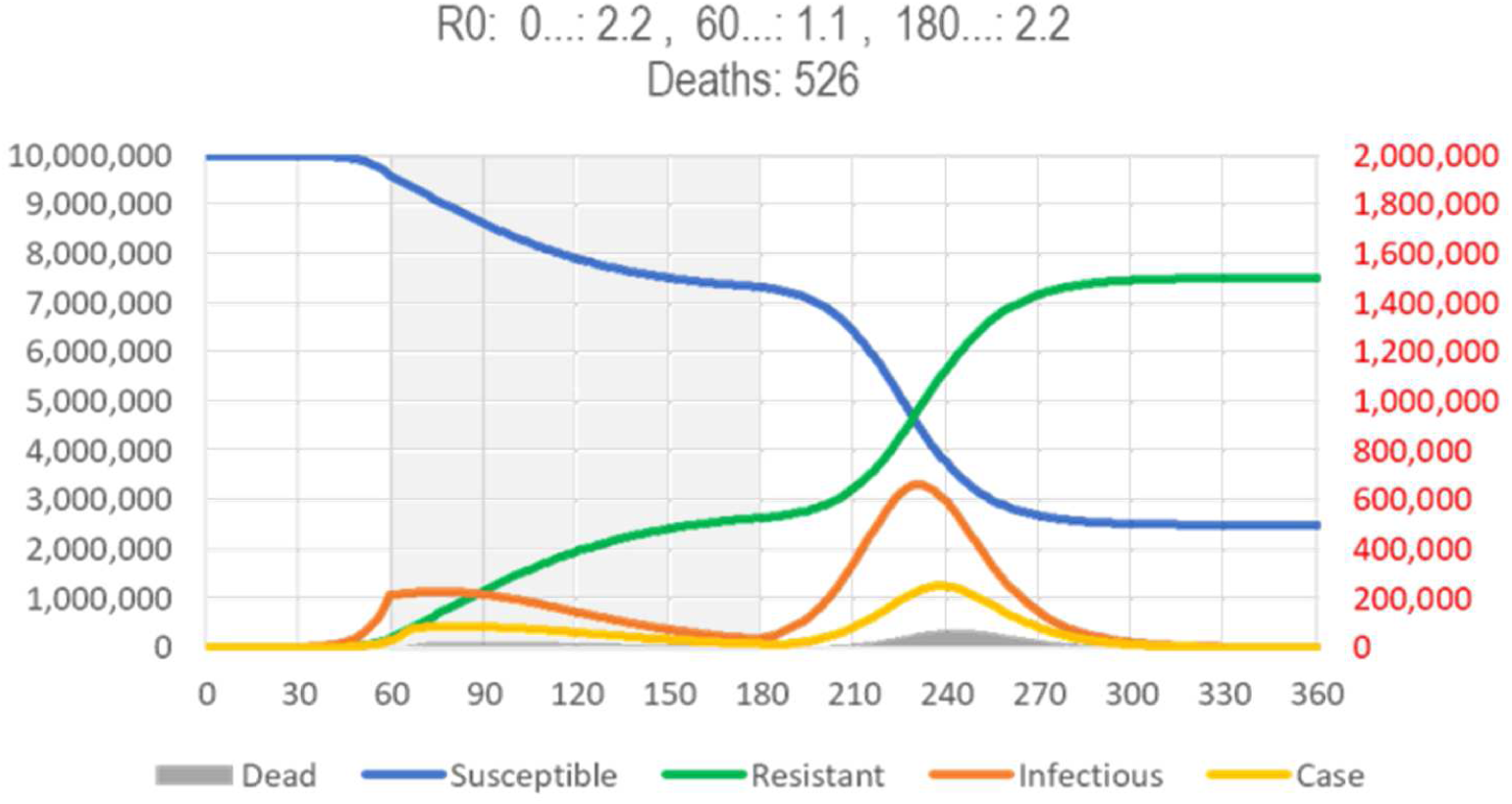
SIR Model of SARS, Effect of Early Lockdown. (see Fig 3 for legend). It is assumed that a highly effective intervention reduces R_0_ by 50% for 4 months, beginning after the appearance of a novel type of cases is noticed. The proportion of symptomatic cases (0.05%/d, i.e., .35% of infected people will become cases and the proportion of cases to die (2%) may change, but the issues discussed here are broadly independent of these assumptions.^(spreadsheet for model calculations available from https://app.box.com/s/pa446z1csxcvfksgi13oohjm3bjg86ql)^

The impact of the interventions discussed on the infections (and, thus, the cases and immunity) depend mostly on the homogeneity of the population, but the number of deaths is highly dependent on how the people respond to the intervention. If parents respond to closure of schools by asking the grandparents to take care of the children, for instance, more of the elderly will get infected and the number of deaths will increase in ways the SIR model cannot predict.

In summary, there is a narrow window-of-opportunity for interventions to “flattening the curve” (reducing R_0_) to be successful in terms of public health:

- Starting after the peak **prevalence** (of infections) has little effect (not shown). The curve goes down, but is not “flattened”.
- Starting at the peak **prevalence** gives the epidemic a “coup de grace”, shortening its duration, albeit at the price of reducing the R/S ratio. The curve is narrower, but also not “flattened” (Fig 12).
- Starting at the peak **incidence** “flattens” (and broadens) the curve and may reduce the number of deaths prevented during the current epidemic, unless
  ○ they increase the proportion of high-risk (elderly) people in the susceptible population at risk of becoming infected (after the children are locked away) or
  ○ they cause behavioral adaptations that increase the contacts of high-risk people with infectious people (such as grandparents taking care of un-schooled children while the parents are working, e.g., in hospitals), but reduces herd immunity and, thus, the chance of another epidemic coming sooner (Fig 13). To prevent a rebound, however, the intervention needs to be extended for several months (Fig 14).
- Starting before the peak **incidence** “flattens the curve”, but also broadens it and causes a rebound, unless the intervention is continued for many more months (Fig 15).

It is herd immunity that stops the spread of an infectious disease, so, in general, one would want to let the epidemic initially run its natural course (or even accelerate it, as people have traditionally done with “measles parties”) to build immunity as fast as possible. If the aim were to reduce the duration of the epidemic and its impact on the economy (and also increase the time until the next epidemic can spread), one would wait until the prevalence of infectious people (I) reaches its peak (in the above model: day 83, red).

Without repeated broad testing, however, that peak prevalence of infections cannot be directly observed, but it is known to be followed about *one week* by peak number of diagnoses (new cases). This is too late to make a decision, but the SIR model shows that the peak in diagnoses is preceded by *two weeks* by the “turning point” in cases where the curve of the new cases changes from increasing ever faster to increasing ever more slowly (day 76). The turning point can be estimated from the observed cases in time to making a decision. (It is also about 50% of the peak number of new cases, which one might be able to predict.) Hence, peak prevalence (of infections) follows the turning point/half peak (in number of cases) by about a week. **The window of opportunity for starting an intervention is the week following the turning point in number of diagnoses (new cases) per day**.

### Responses by country in relation to the turning/decision point

#### Asia

China initially (on 01-23) only closed means of transportation in Wuhan and other Hubei cities, ^[https://www.bbc.com/news/world-asia-china-51217455]^ causing 100,000s to leave the city before, in the afternoon, closing major highways.^[https://www.bbc.com/news/world-asia-china-51217455]^ Family outdoor restrictions were issued on 02-01..05 ^[https://en.wikipedia.org/wiki/2020_Hubei_lockdowns]^ (the week following the 01-31 turning point), non-essential companies were shut down on 02-13, and schools were closed on 02-20, ^[https://en.wik-ipedia.org/wiki/2020_Hubei_lockdowns]^ **two and three weeks, respectively, after the turning point**, when interventions are not effective anymore.

South Korea controlled the epidemic **without imposing a lockdown** on its people^.(https://www.science-mag.org/news/2020/03/coronavirus-cases-have-dropped-sharply-south-korea-whats-secret-its-success)^ and without shutting down its economy, even though the government was initially accused of complacency. ^(https://www.nytimes.com/2020/03/23/world/asia/coronavirus-south-korea-flatten-curve.html)^ People in the city of Daegu, where the Shincheonji Church has its center, were merely asked to “self-quarantine” (“voluntary lockdown”) from 02-23 (the peak of the first wave). ^https://www.newyorker.com/news/news-desk/how-south-korea-lost-control-of-its-coronavirus-out-break)^ On 02-2?,(a day before the peak of the main wave, or only **three days after the turning point**), the KCDC issued a nation-wide recommendation for “social distancing” ^(https://www.institutmon-taigne.org/en/blog/fighting-coronavirus-pandemic-east-asian-responses-republic-korea-mass-testing-targeted)^ Still, the recommended “social distancing” may have prevented herd immunity from developing, as suggested by the continuing low number of cases becoming infected (including some deaths) (Fig 13).

#### Europe

European governments, aiming to take advantage of being forewarned, ordered lockdowns much earlier in the epidemic.

In Italy, the government ordered a nationwide close of restaurants, bars, and most stores more than a week earlier, on 03-11, ^(WSJ, 2020-03-11)^ **four days before the turning point** where the intervention prolongs the epidemic and “social distancing” may cause a rebound (Fig 15). On 03-22, the lockdown in the Italian region of Lombardy was tightened to ban sports and other physical activity, as well as the use of vending machines, an intervention that could have been effective if the virus had spread in the population as a whole, but is unlikely to have changed the dynamic of the epidemic within nursing homes and, especially, to reduce the number of elderly people who died with, but not of the virus.

In Spain, 2020 excess mortality exceeded projections for only four weeks (Fig 16), starting on the same day as the main shutdown (03-14). As infections must have started and peaked about three weeks earlier, the shutdown may have hastened the decline (Fig 12), but couldn’t caused much “flattening” of the curve, which also has the normal, narrow, 4-week width.

**Fig 16:**
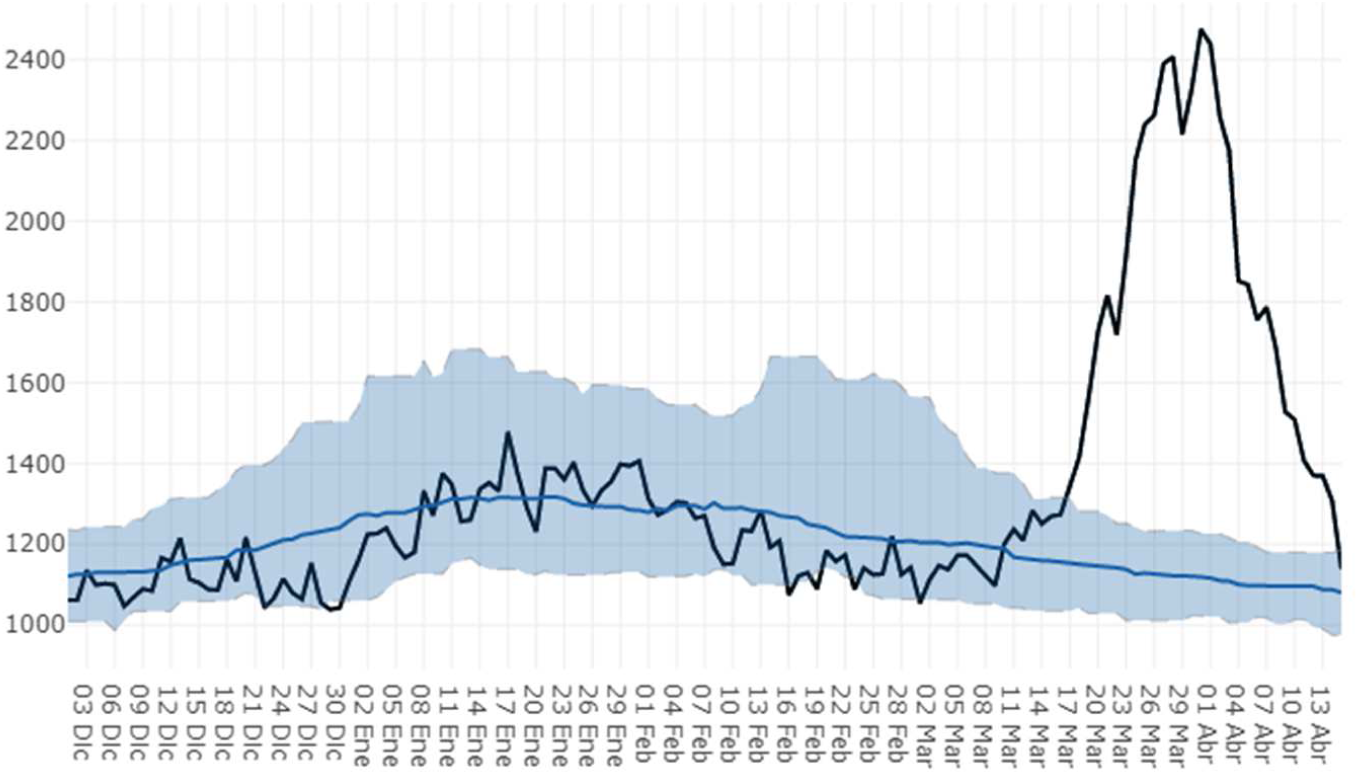
2020 Excess Mortality in Spain. 2020 deaths (black line) vs … estimated deaths (blue line, area: 99% confidence interval) (https://momo.isciii.es/public/momo/dashboard/momo_dashboard.html#nacional %20%3C https://momo.isciii.es/public/momo/dashboard/momo_dashboard.html#nacional%3E)

Germany ordered schools to be closed and various other restrictions on or before 03-16/17, depending on the state ^(https://de.wikipedia.org/wiki/COVID-19-Pandemie_in_Deutschland#Ausgangsbeschr%C3%A4nkungen)^ and a national curfew on 03-22. Taken together, “social distancing” started effectively **several days before the 03-22 turning point**, so that the epidemic is unlikely to reach herd immunity within the near future. Hence the epidemic will not cease for several months, unless restrictions are removed, and, thus, may rebound in autumn (Fig 15). To lessen the rebound, the Academy of Sciences Leopoldina, advised the German government on 04-13 to relax the shutdown, because “our dignified existence in a social, cultural, and also economic way is at stake” and recommended the gradual reopening of some schools and restaurants ^(https://www.dw.com/en/coronavirus-what-is-the-academy-of-sciences-leopoldina-that-advises-the-german-government/a-53115524)^ and, in particular, to reopen primary and early secondary schools “as fast as possible”. ^(https://www.zeit.de/gesellschaft/2020-04/leopoldina-gutachten-coronavirus-schuloeffnungen-abiturienten-infektionsschutz)^ In some states, schools will begin to reopen from 04-20.^(https://www.faz.net/aktuell/ge-sellschaft/gesundheit/coronavirus/leopoldina-raet-abschlussklassen-und-kernfaecher-zuerst-16723112.html)^

Of particular interest is Sweden, where no lockdown was implemented. Similar peak incidence (65–80/M/d), below Europe’s 100/M/d in all Scandinavian countries and a slower increase in incidence in Sweden than in both Norway and Denmark are consistent with the “lockdown” in the latter countries having little, if any effect. Note that differences in lethality are characteristics of the population and, as such, also not affected by a “lockdown” (Fig 9).

In Iceland, which also did not impose a lockdown on its citizens, the epidemic reached a peak at <40/M/d in late March and has since declined to <5/M/d.

#### North America

In the North America, the virus arrived about a week after Europe. Responses varied not only by country, but in the U.S., like in Germany, also by state. An estimated peak of 1.216–4,136 deaths per day in the U.S.^(Murray 2020)^ on 04-16,^(http://www.healthdata.org/covid/updates, accessed on 2020-04-04)^ up from the average of 1,010 on 04-03/04 would be consistent with a turning point about 3 weeks earlier, around 03-27 (Fig 10b).

In NY, one of the epicenters of the U.S. epidemic, “social distancing” started about 03-15 with closure of schools being announced and 03-17 with restaurants being closed, and intensified on 03-22 with the closure of all non-essential businesses (similar actions were taken in New Jersey and Connecticut). Given the fast rise in number of cases in NYC turning on 03-22 (Fig 10c), which was also observed in Detroit, Chicago, and Dallas, see ^[https://www.nytimes.com/interactive/2020/03/27/upshot/corona-virus-new-york-comparison.html, accessed 04-03]^), an intervention at 03-22 started at the turning point (Fig 13). Hence the virus will spread in the population much longer with “social distancing”, and a reduction of deaths through the intervention is possible (albeit not guaranteed). A turning point of 03-22 in NYC is consistent with a projected peak^(Murray 2020)^ of 524–1,090 deaths on 04-10 ^(http://www.healthdata.org/covid/updates, accessed on 2020-04-04)^ compared to the average of 246.5 on 04-02/03.

In California, a shutdown was ordered on 03-19 ^(Executive order N-33-20)^ and many states and cities in the US followed suit, mandating schools, restaurants, and “non-essential” businesses. The turning point in the In the U.S. as a whole, however, cannot have been earlier than 03-27 (Fig 10). Hence, social distancing was ordered before the epidemic reached its turning point, and a “flattened”, but much broader with the virus staying in the population until the intervention is changed (or ignored) so that herd immunity can be reached.

#### Oceania

Other than travel restrictions (14-day quarantine for non-residents and travel restrictions between states, similar to what other countries have enacted) the AU government has not required (although recommended) people to go to work or closed schools,^(https://www.health.gov.au/news/health-alerts/novel-coronavirus-2019-ncov-health-alert/how-to-protect-yourself-and-others-from-coronavirus-covid-19/social-distancing-for-coronavirus-covid-19)^ or financed a bailout to prevent a burndown of the market (as in the U.S.). Instead, it provided funds for ^(https://www.health.gov.au/news/health-alerts/novel-coronavirus-2019-ncov-health-alert/government-response-to-the-covid-19-outbreak, accessed 03-30)^

- delivering an AU$20.7B (AU$800 per person) support package (investment, jobs, health)
- opening fever clinics and funding home delivery of prescription medicines.

## Discussion

### Strengths and shortcomings

A major strength of this analysis of the epidemiological data is that it does not rely on epidemiological models with questionable assumptions. Instead, the results reflect raw incidences over time as reported by the ECDC, depicted by country or region of neighboring countries.

A shortcoming of such an entirely data-based approach is that it lacks the sophistication and potential additional insights that could come from fitting, e.g., differential equation models. The only modeling assumption made is that curves should be “smooth” (except when reporting artifacts are suspected), but even then, data were redistributed only to the directly neighboring day.

Still, the evidence is strong enough to draw qualitative conclusions about possible scenarios for the spread of SARS-CoV-2 in the near future. Also, the results suggest strategies to explore the variability of the SARS-CoV-2 virus strains and to select prevention strategies.

### Epidemiologic evidence for (at least) two different strains of SARS-CoV-2

During the 2003 SARS epidemic the number of new cases peaked about three weeks after the initial increase of cases was noticed and then declined by 90% within a month. Table 1 shows the relevant timepoints for the 2020 COVID-19 epidemic. The SARS-C0V-2 data also suggest that it takes at least a month from the first case entering the country (typically followed by others) for the epidemic to be detected, about three weeks for the number of cases to peak and a month for the epidemic to “resolve”. This data is consistent with the results from the SIR model (see Epidemiological Models).

The 2003 SARS and the 2020 SARS-CoV-2 are not only similar with respect to genetics (79% homology),^(Lu 2020)^ immunology,^(Ahmed 2020)^ involvement of endocytosis (also with influenza and syncytial viruses),^(Behzadi 2019)^ seasonal variation (same season in the northern hemisphere also with influenza, syncytial, and metapneumo viruses)^(Olofsson 2011)^, evolution (origin in bats, 88% homology),^(Benvenuto 2020; Malik 2020)^ but also with respect to the duration between emergence and peak of cases as well as between this peak and resolution of the epidemic (Table 1). Based on these similarities, one could have predicted the natural COVID-19 epidemic to end before 04-15 in Europe and about two weeks later in the U.S., but the interventions to “flatten the curve” seem to have extended the duration.

The time and height of the peak incidence <u>o</u>f cases in the different countries are consistent with the hypothesis that SARS-CoV-2 moved step-by-step westward from China, via other Asian countries to Middle East (Iran. Qatar, and Bahrain), Southern Europe (Italy, followed by its neighbors CH/F/ES/AT/SI), central and northern Europe, and, finally, the US.

Viruses improve their “survival” if they develop strategies to coexist with the (human) host.^(Woolhouse 2007)^ Multiple coronaviruses have been found to coexist in bat populations.^(Ge 2016)^ The emerging COVID-19 data is consistent with the hypothesis that (at least) two SARS-CoV-2 strains have developed. One strain, which traveled through South Korea, remained more infectious, while the other strain, which traveled through other Asian countries lost more of its infectiousness. The strain that passing through South Korea and then Iran and Italy (SKII strain) showed high lethality in Iran and Italy, but less lethality when it traveled to Italy’s neighbors, either because of differences in health systems, because the strain mutated back, or because a strain arriving directly from Asia had the advantage of spreading first. Only sequencing samples from these countries can help to answer these questions. Coronavirus in New York came mainly from Europe.^[https://www.nytimes.com/2020/04/08/science/new-york-coronavirus-cases-europe-genomes.html]^

### Changes in infectivity and lethality between China and Europe

Mainland China is not reporting relevant numbers of new cases anymore and Hubei reports no new cases since 03-19. The number of new cases in South Korea also has declined to low levels since its peak around 02-30. Maritime Southeast Asia continues to show low levels of new cases only (<<u>3</u>/M/d), with the possible exception of Singapore and Malaysia/Brunei. The Philippines are slowly approaching a 3/M/d incidence.

The data are consistent with the same “SKII” strain traveling from China via South Korea and Iran to Italy. Iran was hit about a week after South Korea (around 03-07), with a similar peak incidence, but higher lethality (red bars in Fig 2). The data also suggests a second wave of infections in Iran, with a peak at 15/M/d on 03-15 and a third on 04-01 with a peak of 35/M/d.

Italy was hit a week after the first wave in Iran (which peaked around 03-07/08, Fig 4a). Incidence in Italy reached a substantially higher incidence than reported in Iran. The peak incidence in Italy was reached on 03-22 (at about 100/M/d). Without sufficiently detailed genetic data, it is not clear whether the high lethality in Italy is due to genetic variations in the virus or to Italy having the second oldest populations in the world (after Japan). A 03-20 report by the Istituto Superiore di Sanita,^(COVID-19 Surveillance Group 2020)^ however, implicates that age and comorbidities played a role – among 3200 deaths, mostly in Lombardy and Emilia-Romana, median age was 80 years (IQR 73-85, only were 36 below the age of 50), 98.8% had at least one comorbidity (hypertension: 74%, diabetes: 34%, ischemic heart disease: 30%, atrial fibrillation, 22%, chronic renal failure:

The epidemiological data does not support the hypothesis that SARS-CoV-2 spread from Munich in Germany to Italy.^(Kupferschmidt 2020)^ Instead, the virus may have spread from Italy to its neighboring countries, Switzerland, France, Spain, Austria, and Slovenia, within just a few days of arriving from Iran. The top incidence seems to be less than half of that in Italy and the lethality is lower, too. While Italy has many people 65 years and older (23%, second only to Japan ^data.worldbank.org/indicator/SP.POP.65UP.TO.ZS)^, the relatively small differences or age distribution within Europe (e.g., Germany: 21%) are unlikely to account for much of this difference. A possible explanation (indicated in Fig 6) is that the less virulent strain(s) arriving from other parts of Asia may have had a head start in those countries, so that imported infections from Italy met subjects who had already developed (cross) resistance against both strains.

The parts of Europe directly hit by the SKII strain are already declining from a level of about 100/M/d, the other parts of Europe may have reached their peak of about 90/M/d (but with lower lethality) on 04-04.

### Predictions for COVID-19 in North America

From Table 1, SARS-CoV-2 has arrived in the U.S. almost a week after it arrived in Europe and incidence of COVID-10 in Europe peaked around 04-04. The CDC is monitoring visits to U.S. hospitals for influenza-like illnesses (ILI). Hospital utilization (Fig 18) was highest around week 42 (mid Oct 2009), and week 5 (early Feb, 2018). In 2019-2020, the U.S. was hit by three epidemics: influenza epidemics in late Dec, 2019, and early Feb, 2020, and the COVID-19 epidemic where hospital visits peaked in week 12 (03-16..22), about three weeks before the peak in reported (with some delay) cases around 04-09 (Fig 10) with a substantial drop by week 13 (03-23..29). By 04-05, the number of hospital visits had declined about 67% toward the national baseline.

**Fig 17:**
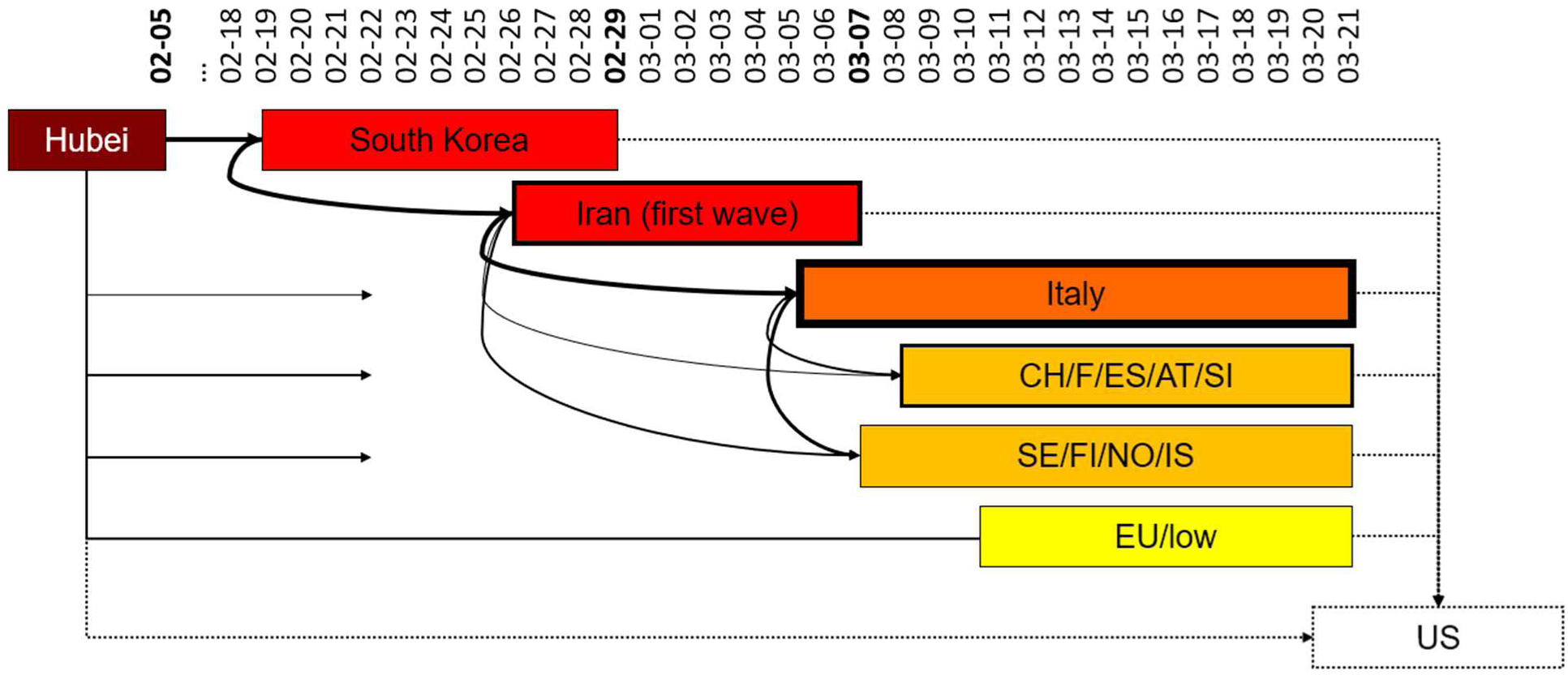
Hypothetical virus transmission pathways. Connection width: number of contacts, box colors: infectiousness, box borders: lethality, dotted connections/borders: unknown. The end date of a box indicates the date of peak incidence, if known (bold date).

**Fig 18:**
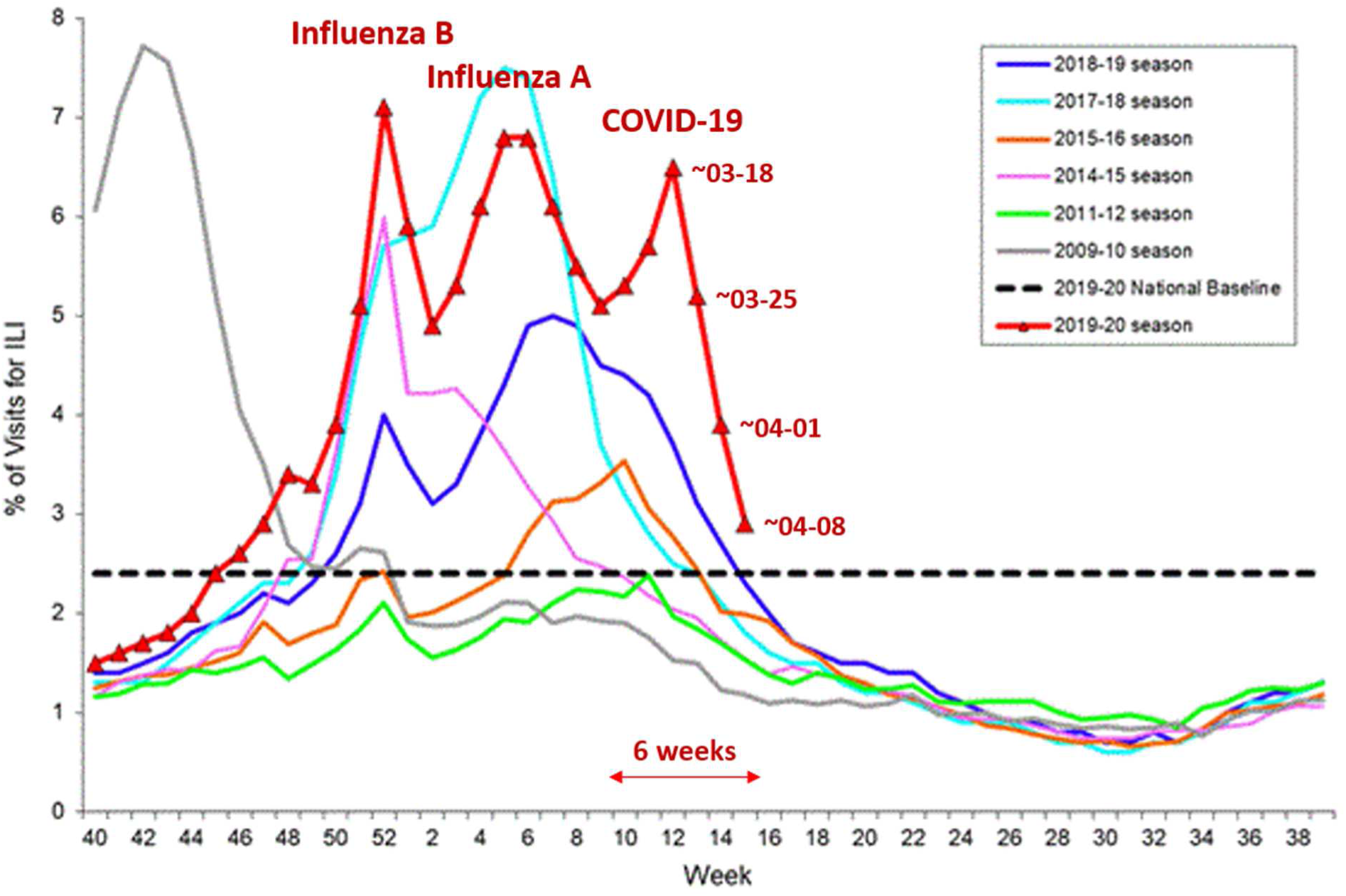
Percentage of US Hospital Visits of Influenza-like Illness (ILI). Reported by the U.S. Outpatient Influenza-like Illness Surveillance Network (ILINet). Weekly national Summary, 2019-2020 and Selected Previous Seasons. The 3^rd^ peak of the 2019-20 season (03-16..22) reflects hospital utilization by COVID-19 patients.^(https://www.cdc.gov/flu/weekly/in-dex.htm, https://www.cdc.gov/flu/weekly/weeklyarchives2019-2020/images/ILI15_small.gif, accessed 2020-04-17f^

U.S. lethality is similar to that of Europe as a whole. Hence, there is no evidence for a high lethality strain dominating the epidemic in the U.S., with the possible exception of NYC. U.S. incidence peaked on 04-05 at about 100/M/d (34,000 cases per day). As in some European countries, the cumulative incidence could reach 4,000/M or a total of 1,300,000 cases, and, at a lethality of about 3%, about 40,000 deaths. Case definitions and reporting guidelines, however, have been changed in the US (e.g., from “death of” to “death with”), so that the number of total deaths may eventually rise to the level of about 60,000, to which the U.S. government has lowered its predictions. A total of 28,000 deaths have already been reported, Even with this broad definition, the number of U.S. deaths over the course of the epidemic would be within the normal range of 16,000–78,000 influenza deaths per season from pneumonia and respiratory/circulatory complications alone, which also occur predominantly among people at 65 years of age and older.^(Rolfes 2018)^. The precise number of people dying will, of course, depend on how easily they can access health care to be treated against severe complications (e.g., pneumonia or respiratory distress).

### A historical perspective

This is not the first, and likely not the last time, that well-intentioned public health policies are inconsistent with our understanding of how epidemics spread. For instance, during much of the HIV epidemic, there was widespread fear that HIV could establish itself in the population as a whole, even though the data (including data showing absence of transmission to the wives of hemophiliacs)^(Wittkowski 1995a)^ and models ^(Wittkowski 1992; Seydel 1994)^ contradicted this fear.^(Wittkowski 1995b; 1996)^ These results have been repeatedly confirmed.^(Centers for Disease Control and Prevention 2019)^; Haddad 2019) In the case of heterosexual transmission of HIV one could argue that there was little risk associated with a the public health policy promoting condom use, but in the case of COVID-19 prevention, ignoring models and data may carry substantial risk.

During the AIDS epidemic, epidemiologists had the advantage that, in addition to the date of report, the date of diagnosis was available for analysis so that variations in reporting delays, such as mid-February in China, 03-20 in Germany, and 03-26 in Canada, could be accounted for. Unfortunately, the public COVID-19 data lacks that information.

### Implications for prevention

In China and South Korea (and the first wave in Iran), incidence peaked after about 2 weeks and then declined. In Europe it took and in the U.S. it takes twice as long. The shorter duration of the epidemic in China and South Korea, however, does not demonstrate the effectiveness of social distancing, because the social distancing started too late to be effective. The longer duration in Europe is consistent with premature interventions (aiming to “flatten the curve”) prolonging the epidemic (“broadening the curve”).

A major problem with respiratory diseases is that one cannot stop all chains of infections within families, friends, neighbors, …. Even after a couple of weeks of “lockdown” there will be a few infectious persons, and as long as there are enough susceptible people in the society, this is enough to re-start the epidemic until there are enough immune people in the society to create “herd immunity”. Hence, one would expect the cases to appear in waves (Fig 15, the period of the “lockdown” corresponds to March to May, 2020 in the U.S.). Such waves of cases have been seen in different countries and the longer than expected duration of the epidemic supports the hypothesis that the social distancing / lockdown interventions had some effect, albeit at a high cost for approx. 10% of deaths saved.

The epidemic of Oceania, together with those of China, South Korea, and Iran are consistent with the model results suggesting that a natural COVID-19 epidemic peaks two weeks after the first cases are seen, and then declines with financial and medical assistance from the government to prevent deaths to reduce the burden on the health system and damage to the economy.

The data from Scandinavia (Fig 9) also does not support the effectiveness of social distancing. The epidemic curves of Sweden and the surrounding Norway, Denmark, and Finland do not indicate that imposed “social distancing” rules in the latter countries, but not in Sweden, had a major impact. Iceland, which, like Sweden, did also not impose a lockdown on its citizens had a particularly mild epidemic.

This analysis of the publicly available data suggests that at the time Italy imposed quarantine on the Lombardy and adjacent regions on 03-08, the SKII virus strain had already reached the adjacent countries (Switzerland, France, Spain, Austria, Slovenia). Even though the lockdown started early (03-08), which may have caused a rebound when compliance declines.

Some containment strategies could even be counterproductive in other ways. For instance, the simple model used in Fig 15 does not account for age-stratification. In diseases such as COVID-19, children develop mostly mild forms, elderly people have a high risk of dying.^(Zimmermann Curtis 2020)^ Hence, containment of high-risk groups, like elderly people in nursing homes (see the Washington State example) is highly effective in protecting them from becoming infected and in reducing the pool of children and young adults that would have to be infected to reach herd immunity. A substantial increase in the duration of the epidemic by preventing immunity to develop among the young, however, might make effective containment of the elderly more difficult and, thus, increase the number of deaths among the elderly.

## Conclusions

Until a vaccine will become available, the only pharmacological strategy to reduce the number of deaths is to reduce the damage the infection (and immune system) does, e.g., by reducing the initial viral load,^(Chu 2004)^ and making sure that people get treated at the earliest signs of pneumonia.

Aside from separating susceptible populations (elderly and high-risk subjects, e.g., in nursing homes) from the epidemic, which is effective as long as virus is circulating, public health intervention aiming to contain a respiratory disease need to start within a narrow window of opportunity starting when or a week after the curve of the new cases changes from increasing faster to increasing more slowly. Only if stopping the epidemic from generating a sufficient number of immune people is avoided can containment efforts stop after about a month or two (depending on late or early start, respectively), when the ratio of infectious vs immune people is low enough for preventing the disease from rebounding. When the window of opportunity has been missed, any type of containment has only limited impact on the course of the epidemic, but high impact on economy and society.

So far, the hospital systems in most affected countries have shown to be able to handle the COVID-19 epidemic. As epidemics in maritime Asia and the southern hemisphere remain a possibility and some develop within the next weeks, the use of data and scientifically sound models may help to time interventions to optimize their effect. In particular of the risk of prolonging the epidemic and increasing the number of deaths by starting interventions (“flattening the curve”) before the turning point in the number of cases is reached.

To determine that time point, case data collected and reported needs to contain not only the date of report, but also the date of “diagnosis” and whether the patient had clinical symptoms or was merely tested positive and whether the patient was positive for circulating virus RNA/DNA (currently infectious) or antibodies (already immune).

## Data Availability

The data is publicly available

http://ecdc.europa.eu

## Notes

### Competing Interest Statement

Dr. Wittkowski is currently the CEO of ASDERA LLC, a company discovering novel treatments for complex diseases from data of genome-wide association studies. One of these treatments could potentially be effective against virus (including coronavirus) diseases.

### Funding Statement

none

